# Unravelling the molecular mechanisms causal to type 2 diabetes across global populations and disease-relevant tissues

**DOI:** 10.1101/2025.05.05.25326880

**Authors:** Ozvan Bocher, Ana Luiza Arruda, Satoshi Yoshiji, Chi Zhao, Chen-Yang Su, Xianyong Yin, Davis Cammann, Henry J. Taylor, Jingchun Chen, Ken Suzuki, Ravi Mandla, Alicia Huerta-Chagoya, Ta-Yu Yang, Fumihiko Matsuda, Josep M. Mercader, Jason Flannick, James B. Meigs, Alexis C. Wood, Marijana Vujkovic, Benjamin F. Voight, Cassandra N. Spracklen, Jerome I. Rotter, Andrew P. Morris, Eleftheria Zeggini

## Abstract

Type 2 diabetes (T2D) is a prevalent disease that arises from complex molecular mechanisms. Here, we leverage T2D multi-ancestry genetic associations to identify causal molecular mechanisms in an ancestry- and tissue-aware manner. Using two-sample Mendelian Randomization corroborated by colocalization across four global ancestries, we analyze 20,307 gene and 1,630 protein expression levels using blood-derived *cis*-quantitative trait loci (QTLs). We detect causal effects of genetically predicted levels of 335 genes and 46 proteins on T2D risk, with 16.4% and 50% replication in independent cohorts, respectively. Using gene expression cis-QTLs derived from seven T2D-relevant tissues, we identify causal links between the expression of 676 genes and T2D risk, including novel associations such as *CPXM1*, *PTGES2* and *FAM20B*. Causal effects are mostly shared across ancestries, but highly heterogeneous across tissues. Our findings provide insights in cross-ancestry and tissue-informed multi-omics causal inference analysis approaches and demonstrate their power in uncovering molecular processes driving T2D.

## Introduction

Type 2 diabetes (T2D) is a prevalent, complex disease. Large-scale genome-wide association study (GWAS) meta-analyses^1,2^ have advanced the understanding of its underlying genetic architecture. As T2D is a heterogeneous disease^3^ with different underlying pathology between individuals, identifying different causal biological mechanisms can facilitate potential leads for drug development^4^ and interventions in patient populations that maximize clinical benefit. While causal inference is challenging, statistical inference methods such as Mendelian randomization (MR) can estimate the putative causal effect of an exposure on an outcome when specific assumptions are met, using genetic variants predictive of the exposure as instrumental variables (IVs)^5^. Using genetic variation as IVs for molecular phenotypes, i.e. quantitative trait loci or ‘QTLs’, MR has also been successfully employed to provide evidence of causal effects of methylation^6,7^, gene expression^8^, protein^9^ or metabolite levels^10,11^ on T2D risk. However, most of these studies have been performed using blood levels of the putative causal risk factors and have not addressed mechanisms in disease-relevant tissues underlying genetic associations^12^. Furthermore, the majority of studies have focused on populations genetically similar to Europeans (EUR; based on the 1000 Genomes Project phase 3^13^ individuals sampled from continental Europe as a reference), which limits inference of causal molecular mechanisms across diverse global populations. In addition to investigating how causal effects are shared across populations throughout the world, assessing QTLs in these groups facilitates the assessment of molecular traits for which IVs are not available in EUR^14^.

To expand diversity in T2D genetic studies, and improve the generalizability of findings, the T2D Global Genomics Initiative (T2DGGI) has performed a multi-ancestry GWAS meta-analysis gathering genome-wide data from over 2,5 million individuals, including over 700,000 individuals of non-European ancestry15 . Mandla et al16. integrated these T2DGGI data with large-scale omics, identifying evidence of colocalization17 between molecular traits and 56% of the 1,289 T2D GWAS meta-analysis index variants and highlighting the benefit of interrogating data from under-represented global populations. Here, we study the causal links of gene expression and protein abundance levels with T2D risk using MR approaches based on *cis*-QTLs, predicted to have the strongest biological impact on molecular traits18 . We expand previous causal inference studies by (1) leveraging the T2D genetic associations reported by T2DGGI and QTL maps from global populations in single-ancestry analyses and multi-ancestry meta-analyses and (2) investigating causal effects in seven further tissues relevant to T2D. To corroborate our findings, we perform colocalization using a method suitable for polygenic traits, such as T2D, as it allows for multiple causal variants at each interrogated genetic locus.

## Results

MR analyses were performed using quantitative trait loci (QTL) as IVs. Throughout this manuscript, we refer to ‘genes’ for gene expression levels genetically predicted from *cis*-expression QTL (eQTL) and to ‘proteins’ for protein levels genetically predicted from *>cis*-protein QTL (pQTL). We use ‘molecular traits’ to refer to genes and proteins identified in any MR analysis.

### Study design overview

We performed blood MR analyses by using IVs defined from blood eQTL and plasma pQTL from multiple cohorts (**Figure 1**, Supplemental Table 1). Analyses were conducted in an ancestry-aware manner, i.e. within ancestry groups genetically similar to Europeans (EUR), Africans (AFR), admixed Americans (AMR) and East-Asians (EAS) (**Figure 1**). As previously recommended for *cis*-MR analyses^19^, effects were considered as causal if they presented FDR-adjusted p-values (q-values) from the inverse variance weighted (IVW) method lower than 5% with concordant effect across sensitivity analyses, showed no evidence of heterogeneity and pleiotropy, and were corroborated by evidence of colocalization (Methods). Replication was performed in matched genetic ancestry groups depending on data availability (**Figure 1**). We meta-analyzed MR results across genetic ancestry groups, where possible, using random effect models for either genes or proteins, separately, and considered causal effects with a q-value lower than 5% and compelling evidence in at least one cohort (**Methods**). Finally, we performed *cis*-eQTL MR analysis in seven additional T2D-relevant tissues (whole pancreas, pancreatic islets, brain hypothalamus, visceral adipose, subcutaneous adipose, liver, and skeletal muscle). We report causal estimates as odds ratios (OR) per unit of genetically predicted gene expression and protein levels on T2D risk.

**Figure 1:**
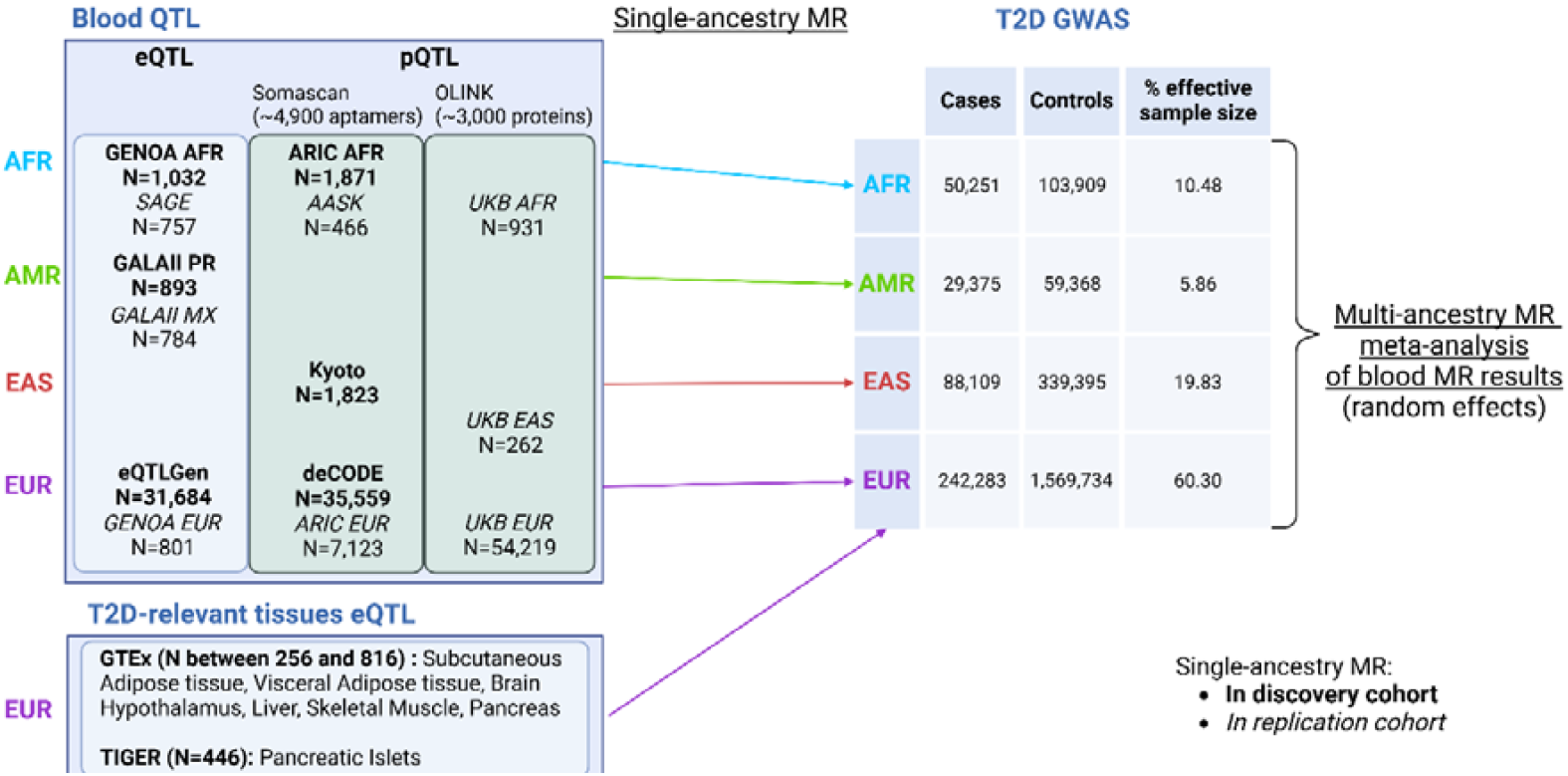
Overview of the cohorts and tissues used to perform single-ancestry MR analyses in populations genetically similar to Europeans (EUR), Africans (AFR), admixed Americans (AMR), and East-Asians (EAS) based on the 1000 Genome Project phase 313 . Discovery cohorts are indicated in bold, and replication cohorts for blood MR analyses in italic. Figure generated with Biorender.

### Low ancestry-related heterogeneity for blood molecular traits causal to T2D risk

We defined IVs in blood for a total of 20,307 genes and 1,630 proteins. A total of 13,878 genes and 1,094 proteins had IVs in at least two genetic ancestry groups and were meta-analyzed using random effect models to obtain a cross-ancestry causal effect estimate. We identify causal effects of expression levels of 81 genes and 5 unique proteins on T2D risk in the MR meta-analysis across genetic ancestry groups (**Figure 2**, Supplemental Tables 2 and 3). Most of these causal effects of genes (93.8%) and proteins (80%) were also significant in the EUR single-ancestry MR analysis, highlighting the driving power of this genetic ancestry group in the meta-analysis, linked to the larger sample size of the corresponding QTL and GWAS datasets. The meta-analysis identified four genes not detected in any single-ancestry MR analyses, highlighting the benefits of combining MR results from multiple genetic ancestry groups (Supplemental Figure 1).

**Figure 2:**
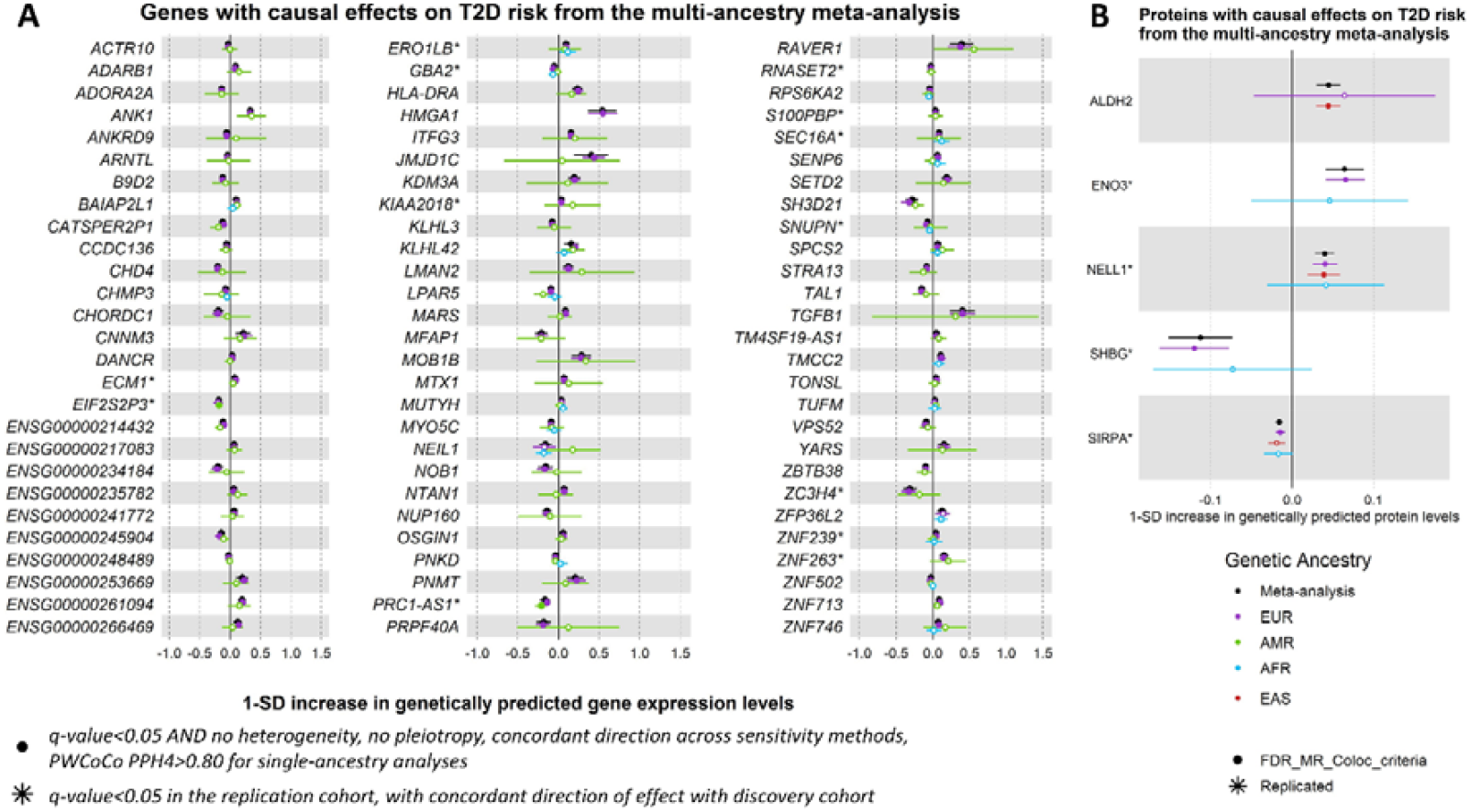
Genes and proteins with causal effects identified in the MR multi-ancestry meta-analysis. Causal estimates from the single-ancestry MR in the discovery cohorts are also depicted. Filled dots represent causal estimates from MR analyses that have a q-value<0.05, and (1) pass the sensitivity criteria and show evidence of colocalization (PPH4>0.8) in single-ancestry analyses, or (2) present nominal significance and meet criteria (1) in at least one cohort entering the meta-analysis. Genes and proteins with causal effects identified in single-ancestry analyses and replicated in independent cohorts from the same genetic ancestry group are denoted with a star. We report causal estimates as odds ratios (OR) for T2D per standard deviation (SD) change in genetically predicted gene expression or protein levels.

Of the 81 genes with significant causal effects on T2D risk in the meta-analysis, evidence of ancestry-related heterogeneity, as determined by a nominal significant Cochran’s Q statistic, was found for only one gene, *KLHL42* (Cochran’s p-value = 3.96×10^2^), and for none of the proteins. Altogether, we find low levels of ancestry-related heterogeneity of causal effects of gene and protein expression levels on T2D risk. However, this observation needs to be confirmed in future studies once equivalent statistical power is achieved across genetic ancestry groups. Many efforts will be required to meet this goal, as most molecular QTL studies are typically underrepresented in non-EUR genetic ancestry groups^20^.

### Assessing QTLs in global populations improves the detection of causal effects

Causal effects of 181 genes were significant in at least one single-ancestry MR analysis, but not in the cross-ancestry meta-analysis, of which 168 (93%) were found in the EUR analysis. A total of 14 genes showed significant causal effects only in non-EUR (nine in AMR and five in AFR), of which nine (69%) were replicated in independent cohorts from matched genetic ancestry groups, strengthening the evidence for their potential causal role on T2D risk (**Figure 3A**, Supplemental Tables 4 and 5). Two examples include *TOLLIP-AS1* in AMR (OR=0.87, q-value=2.25×10^4^) and *PTGES2* in AFR (OR=0.90, q-value=2.40×10^2^), with protective effects against T2D risk (Supplemental Figure 2A-B). When investigating the allele frequencies and effect sizes of the IVs from these two genes, we observed different patterns across genetic ancestry groups. The allele frequencies of the IVs used for *PTGES2* across AFR and EUR were similar, while the effect sizes on gene expression were higher in AFR than in EUR. Conversely, for *TOLLIP-AS1*, the effect sizes on gene expression were of the same magnitude among AMR and EUR, but the allele frequencies of the IVs were higher in AMR than in EUR (Supplemental Figure 2C). Through the examples of *PTGES2* and *TOLLIP-AS1*, we highlight that causal effects observed in only one genetic ancestry group might be related to differences in statistical power rather than causal effects truly absent from other genetic ancestry groups, underscoring the challenge of assuming ancestry-specific causal effects. These differences in statistical power further influence the meta-analysis results (in the meta-analysis: OR=0.92, q-value=0.32 for *PTGES2*; OR=0.94, q-value=0.83 for *TOLLIP-AS1*).

**Figure 3:**
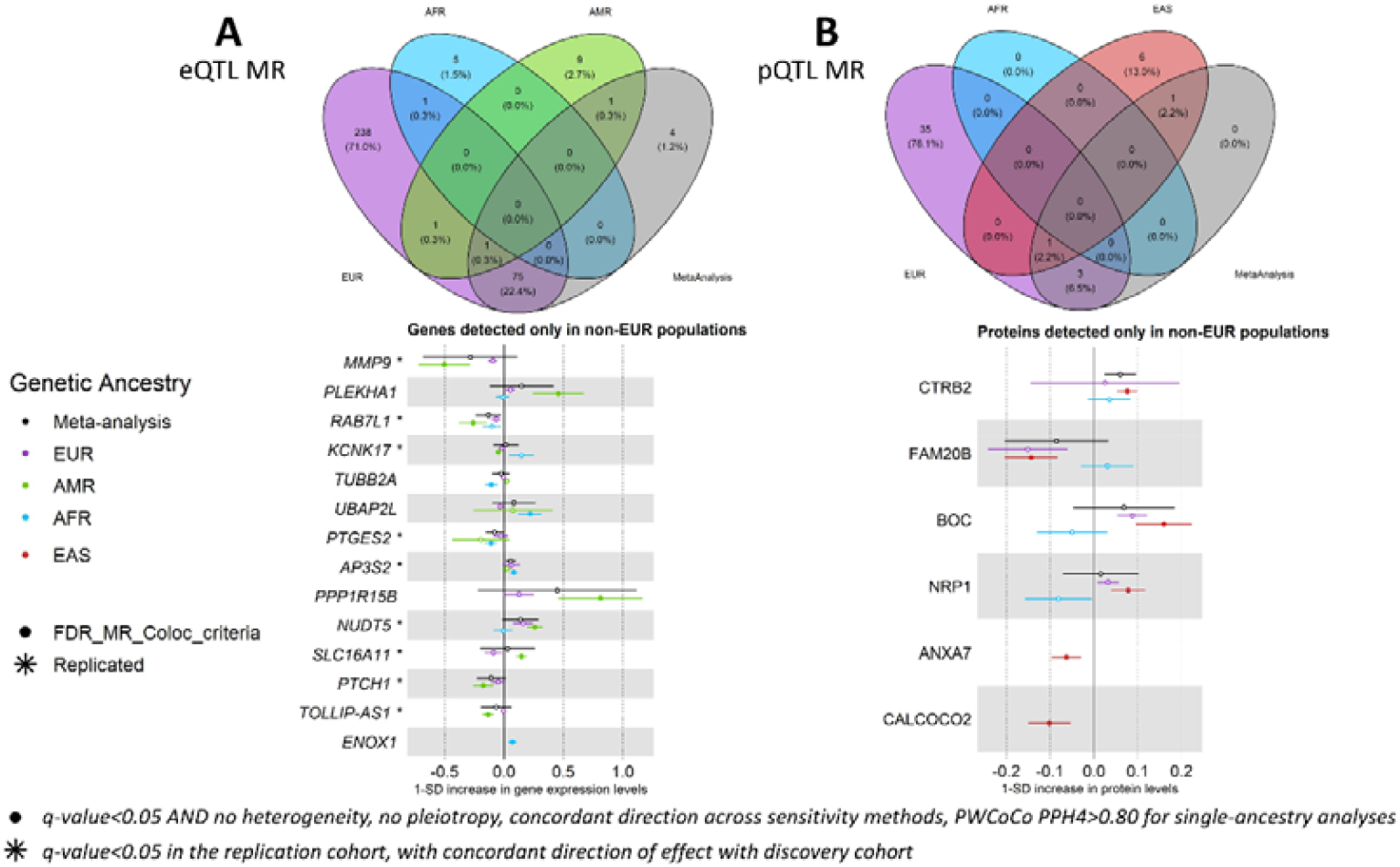
Venn-diagrams showing the overlap of putative causal effects between genetic ancestry groups and forest plots of causal effects of genes (A) and proteins (B) detected only in non-EUR. Filled dots represent causal estimates from MR analyses that have a q-value<0.05, and (1) pass the sensitivity criteria and show evidence of colocalization (PPH4>0.8) in single-ancestry analyses, or (2) present nominal significance and meet criteria (1) in at least one cohort entering the meta-analysis. Genes and proteins with causal effects identified in single-ancestry analyses and replicated in independent cohorts from the same genetic ancestry group are denoted with a star. We report causal estimates as odds ratios (OR) for T2D per standard deviation (SD) change in genetically predicted gene expression or protein levels.

Similarly, there were 27 proteins with causal effects detected in single-ancestry MR analyses but not in the cross-ancestry meta-analysis, of which 23 (89%) were only detected in EUR. The remaining four proteins with significant causal effects only in non-EUR were detected in EAS (**Figure 3B**): CTRB2 (OR=1.08, q-value=3.60×10^8^), FAM20B (OR=0.87, q-value=3.80×10^4^), BOC (OR=1.17, q-value=2.17×10^4^) and NRP1 (OR=1.08, q-value=6.59×10^3^). These four proteins could not be tested for replication in EAS due to the lack of a replication cohort with Somascan proteomics data, and the lack of IVs for these proteins in the EAS UK Biobank^21^ (UKB) (Supplemental Table 6). All four proteins were tested in all available genetic ancestry groups (EUR, AFR, EAS) and presented causal estimates in EUR close to the estimates in EAS. For example, BOC and FAM20B presented causal effects with a q-value lower than 5% in the EUR MR analyses but did not show evidence of colocalization in this ancestry group (BOC: PPH4=0.068 in EUR vs PPH4=0.919 in EAS; FAM20B: PPH4=3.2×10^5^ in EUR vs PPH4=0.974 in EAS). The EUR pQTL dataset used in our study comes from deCODE, an Icelandic population dataset. The lack of colocalization in EUR for these proteins may be due to an LD mismatch between Icelandic pQTL data and the EUR GWAS meta-analysis from T2DGGI. For EAS, colocalization was identified possibly due to a more similar LD pattern between the pQTL and EAS GWAS meta-analysis from T2DGGI (Supplemental Figure 3).

Our results further reveal the benefits of investigating non-EUR QTLs given the increased number of genes and proteins for which IVs are available only in non-EUR. Here, a total of 6,431 genes and 570 proteins were tested only in one genetic ancestry group, of which 3,648 genes (56.7×0025;) and 302 proteins (53.0×0025;) were only tested in non-EUR. Most of these IVs show minor allele frequencies lower in EUR than in non-EUR, especially when considering IVs of genes and proteins tested only in AFR (**Figure 4**). It would therefore require larger sample sizes in EUR QTL studies to detect these IVs, given the expected homogeneous effect sizes across genetic ancestry groups^22^. Examples include *ENOX1* (OR=1.07, q-value=3.80×10^2^ in AFR eQTL MR), ANXA7 (OR=0.94, q-value=1.16×10^2^ in EAS pQTL MR) and CALCOCO2 (OR=0.90, q-value=3.61×10^3^ in EAS pQTL MR). *CALCOCO2* is a gene previously suggested to be associated with T2D^23^, with a knockdown decreasing insulin content in the human pancreatic beta cell line EndoC-βH1^24^. Here, we corroborate this with evidence of a protective effect of increased expression levels are CALCOCO2 against T2D risk.

**Figure 4:**
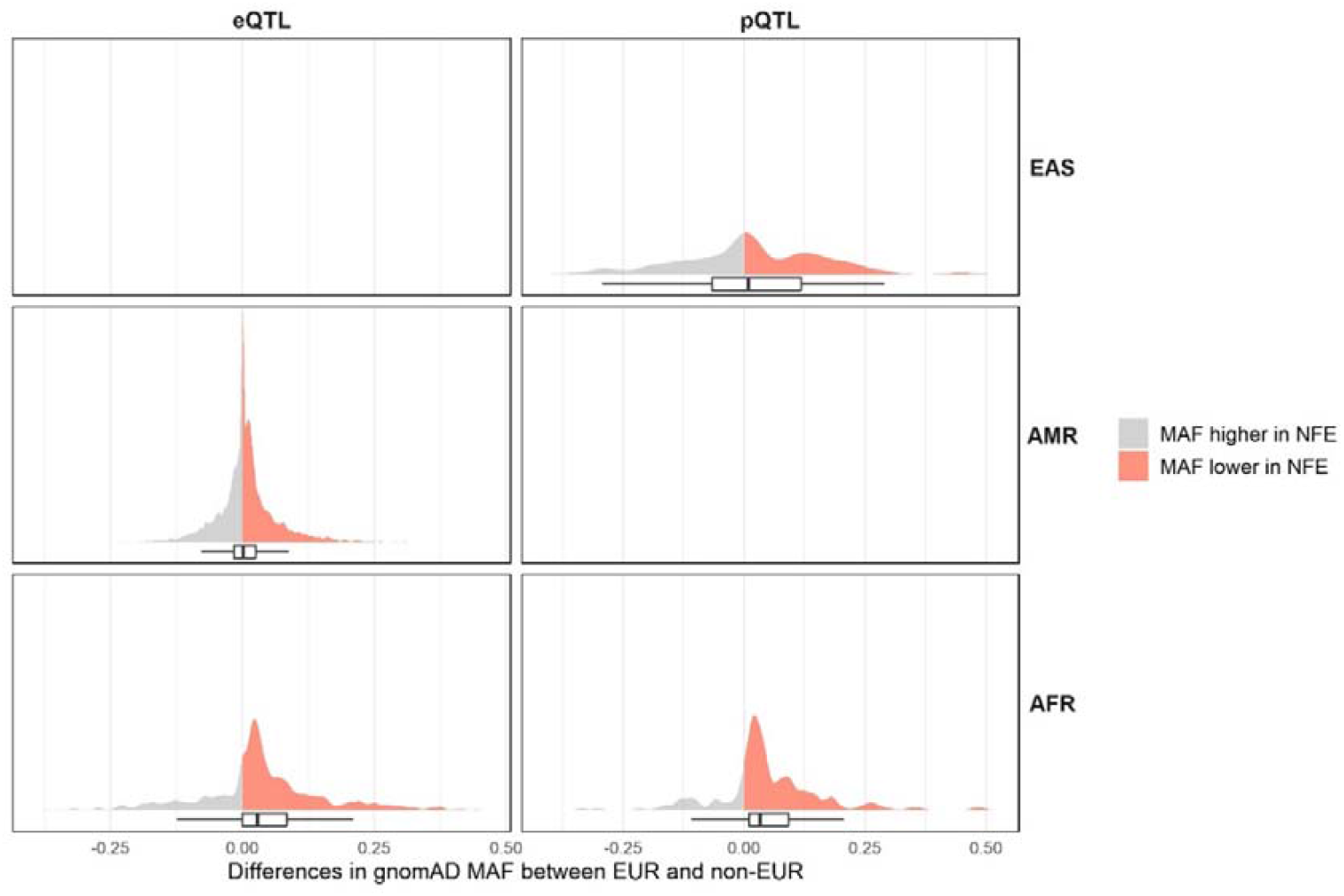
Distribution of differences between EUR and non-EUR in minor allele frequencies (MAF) of IVs for genes and proteins only tested in non-EUR. The differences were computed as MAF_EUR_ -MAF_non-EUR_ . MAF were obtained from the Genome Aggregation Database (gnomAD)25 . gnomADg_NFE_MAF refers to the MAF observed in gnomAD genomes in the Non-Finnish European (NFE) population. Positive differences, i.e. IVs for which the MAF is higher in NFE than in the corresponding non-EUR ancestry group, are represented in red, and negative differences in grey.

### Causal effects present high tissue-related heterogeneity

In addition to blood, MR analyses were conducted in EUR in seven tissues relevant to T2D: subcutaneous adipose, visceral adipose, brain hypothalamus, liver, skeletal muscle, whole pancreas, and pancreatic islets. Causal effects on T2D risk were identified for 70 to 243 genes, depending on the tissue (**Figure 5A**, Supplemental Table 7). Because molecular QTL data in blood are more widely available across multiple genetic ancestry groups and with higher sample sizes than in other tissues, we compared findings from non-blood tissues to the 335 genes identified in any blood eQTL MR analyses. A total of 923 genes showed significant causal effects on T2D risk in at least one T2D-relevant tissue and/or in blood. We observed the largest causal effects in blood, followed by visceral adipose tissue, skeletal muscle, and subcutaneous adipose tissue (Supplemental Figure 4).

**Figure 5:**
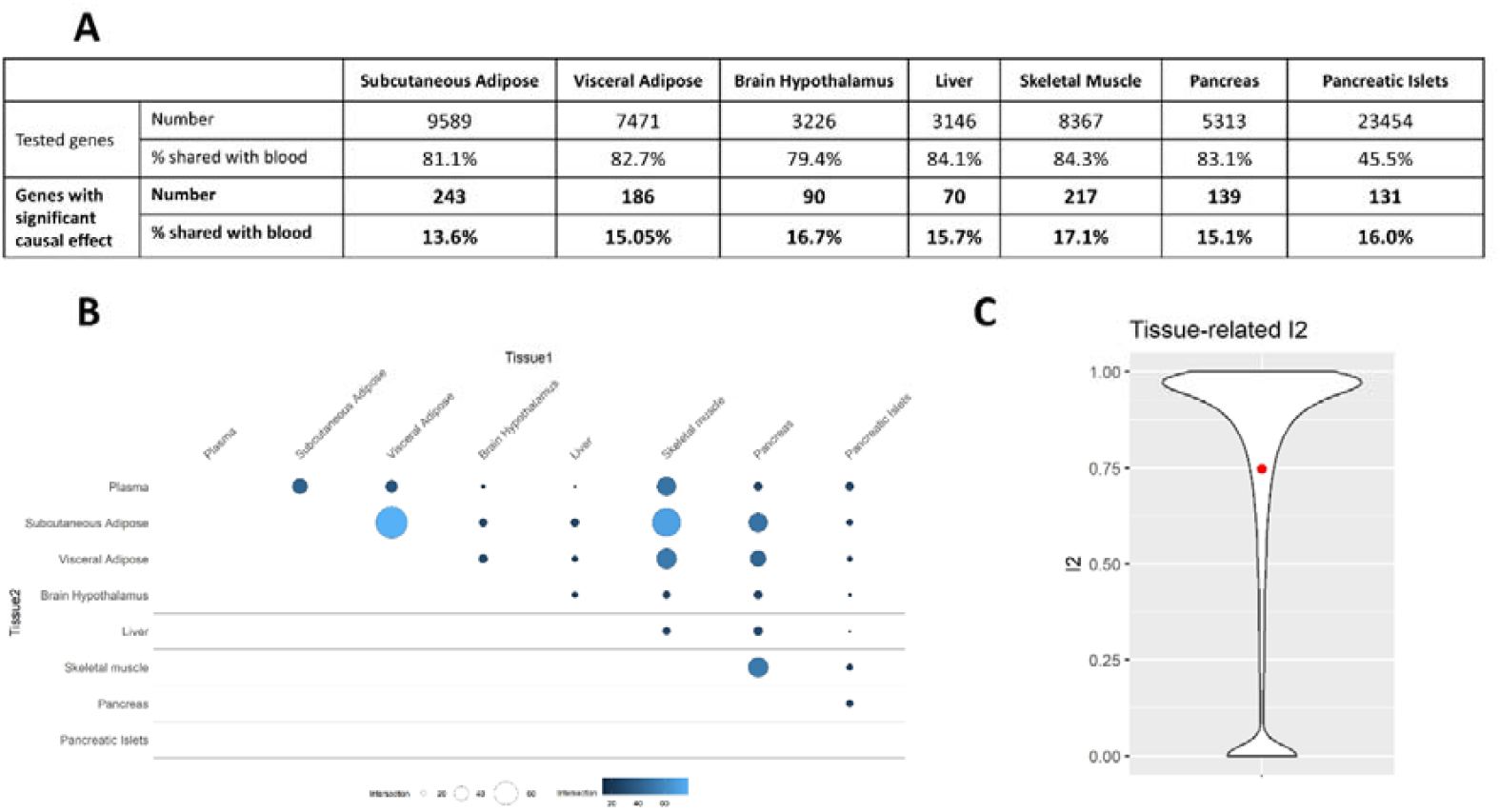
Overview of the results from the MR analyses in T2D-relevant tissues. (A) Number of genes tested in MR analyses from T2D-relevant tissues, with significant causal effects on T2D risk, and percentage of causal effects also detected in blood eQTL MR. (B) Pairwise overlap of significant causal effects across T2D-relevant tissues and blood eQTL MR. (C) Distribution of I2 values representing the heterogeneity of causal estimates for genes tested in at least two tissues (including blood).

We report 328 genes (36%) with significant causal effects in at least two tissues (**Figure 5B**). Only 18% of the genes with causal effects identified in a T2D primary tissue also showed significant causal effects in blood, highlighting the additional information provided by eQTL MR analyses in non-blood tissues. The highest overlap was observed between subcutaneous and visceral adipose tissues, as well as between subcutaneous adipose tissue and skeletal muscle. We observed low overlap between whole pancreas and pancreatic islets, underscoring the importance of investigating fine-scale tissue molecular profiles. Only 28 out of the 328 genes with causal effects on T2D risk in multiple tissues showed concordant directions of effect across tissues. Accordingly, we observed a large tissue-related heterogeneity, as highlighted by the high I^2^ values depicted in **Figure 5C**, with 75% of the genes tested in at least two tissues (14,304 out of 19,020) showing a nominal significant heterogeneity. This tissue-related heterogeneity could be underestimated given the lack of shared IVs across tissues when investigating *cis*-eQTL^26^, with 11,707 genes (21%) in our analyses being tested in only one non-blood tissue. Evaluating the causal effects of all genes in disease-relevant tissues will be needed to comprehensively capture tissue-related heterogeneity and characterize shared and tissue-specific causal mechanisms.

### eQTL and pQTL analyses offer complementary and non-redundant insights

To assess the complementarity between eQTL and pQTL MR findings, we compared the genes and proteins with significant causal effects identified by each analysis (**Table 1**).

**Table 1:**
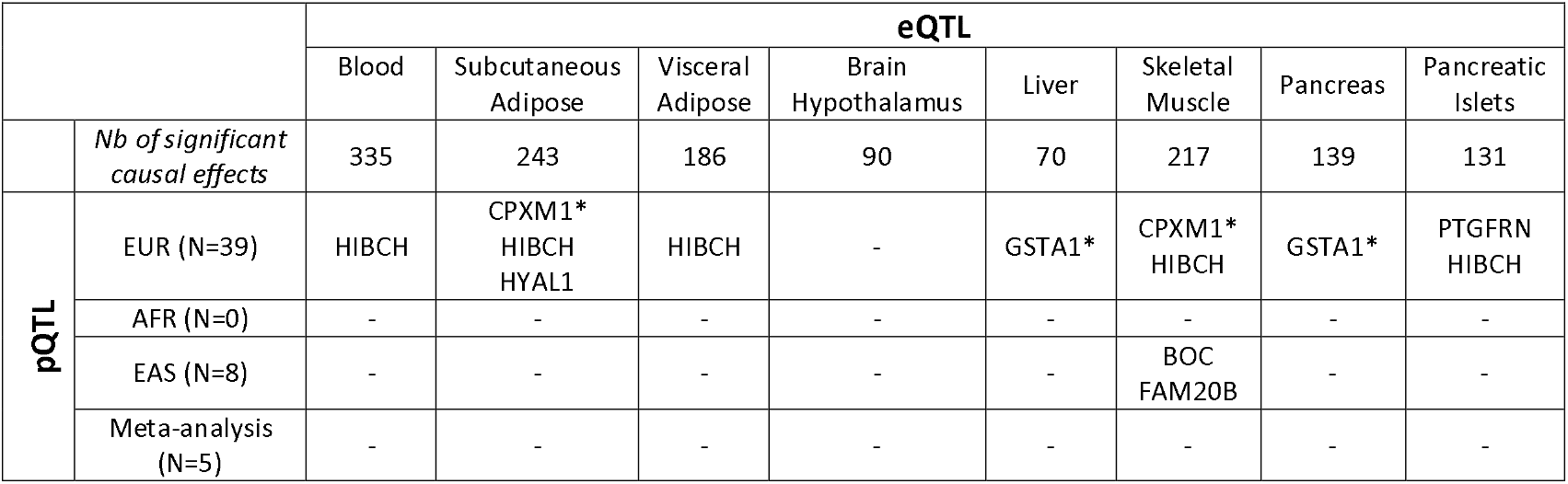
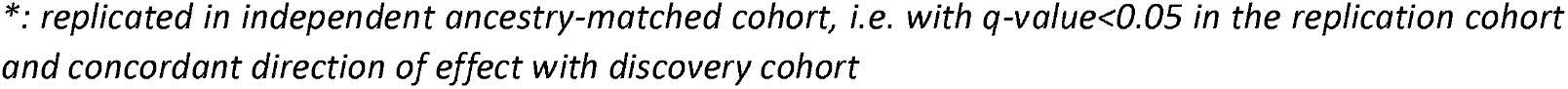
Molecular traits (genes/proteins) with evidence of causal effects on T2D risk from eQTL and pQTL MR. Molecular traits with causal effects replicated in independent cohorts are indicated with a star.

|  |  | eQTL |  |  |  |  |  |  |  |
| --- | --- | --- | --- | --- | --- | --- | --- | --- | --- |
|  |  | Blood | Subcutaneous Adipose | Visceral Adipose | Brain Hypothalamus | Liver | Skeletal Muscle | Pancreas | Pancreatic Islets |
|  | Nb of significant causal effects | 335 | 243 | 186 | 90 | 70 | 217 | 139 | 131 |
| pQTL | EUR (N=39) | HIBCH | CPXM1*<br>HIBCH<br>HYAL1 | HIBCH | - | GSTA1* | CPXM1*<br>HIBCH | GSTA1* | PTGFRN<br>HIBCH |
|  | AFR (N=0) | - | - | - | - | - | - | - | - |
|  | EAS (N=8) | - | - | - | - | - | BOC<br>FAM20B | - | - |
|  | Meta-analysis (N=5) | - | - | - | - | - | - | - | - |

\*: replicated in independent ancestry-matched cohort, i.e. with $q\text{-value} < 0.05$ in the replication cohort and concordant direction of effect with discovery cohort

The absolute number of molecular traits with overlapping evidence of causal effects of gene expression and protein abundance levels was small (seven out of 1,563 molecular traits tested in both eQTL and pQTL MR analyses), in line with the expected complementary information captured by the two molecular levels^27^. Only *HIBCH* was identified in eQTL- and pQTL-based causal inference analyses in blood (eQTL OR=0.96, q-value=1.37×10^2^; pQTL OR=0.95, q-value=4.94×10^4^). Increased expression levels of this gene in pancreatic islets (OR=0.98, q-value=2.9e-3) and visceral adipose tissue (OR=0.96, q-value=1.15×10^3^) also showed a significant protective causal effect against T2D risk (**Figure 6A**). In contrast, higher expression levels of *HIBCH* in subcutaneous adipose tissue (OR=1.06, q-value=5.68×10^4^) and skeletal muscle (OR=1.04, q-value=5.31×10^4^) were causally associated with increased T2D risk. These results complement previous findings where causal effects of *HIBCH* have been reported in EUR from blood eQTL and pQTL analyses^28^, as well as in the liver, pancreas, and visceral adipose tissue^29^. Similarly, increased protein levels of GSTA1 were previously found to be causally associated with increased T2D risk using plasma pQTL MR^30^, and liver tissue *cis*-eQTLs of this gene were found to colocalize with T2D index variants ^16^. Here, we describe concordant results for GSTA1 protein abundance levels from the blood pQTL MR (OR=1.06, q-value=7.49×10^7^) and additionally identify protective effects of gene expression in the liver (OR=0.92, q-value=7.78×10^6^) and in whole pancreas (OR=0.95, q-value=1.37×10^3^) (**Figure 6B**).

**Figure 6:**
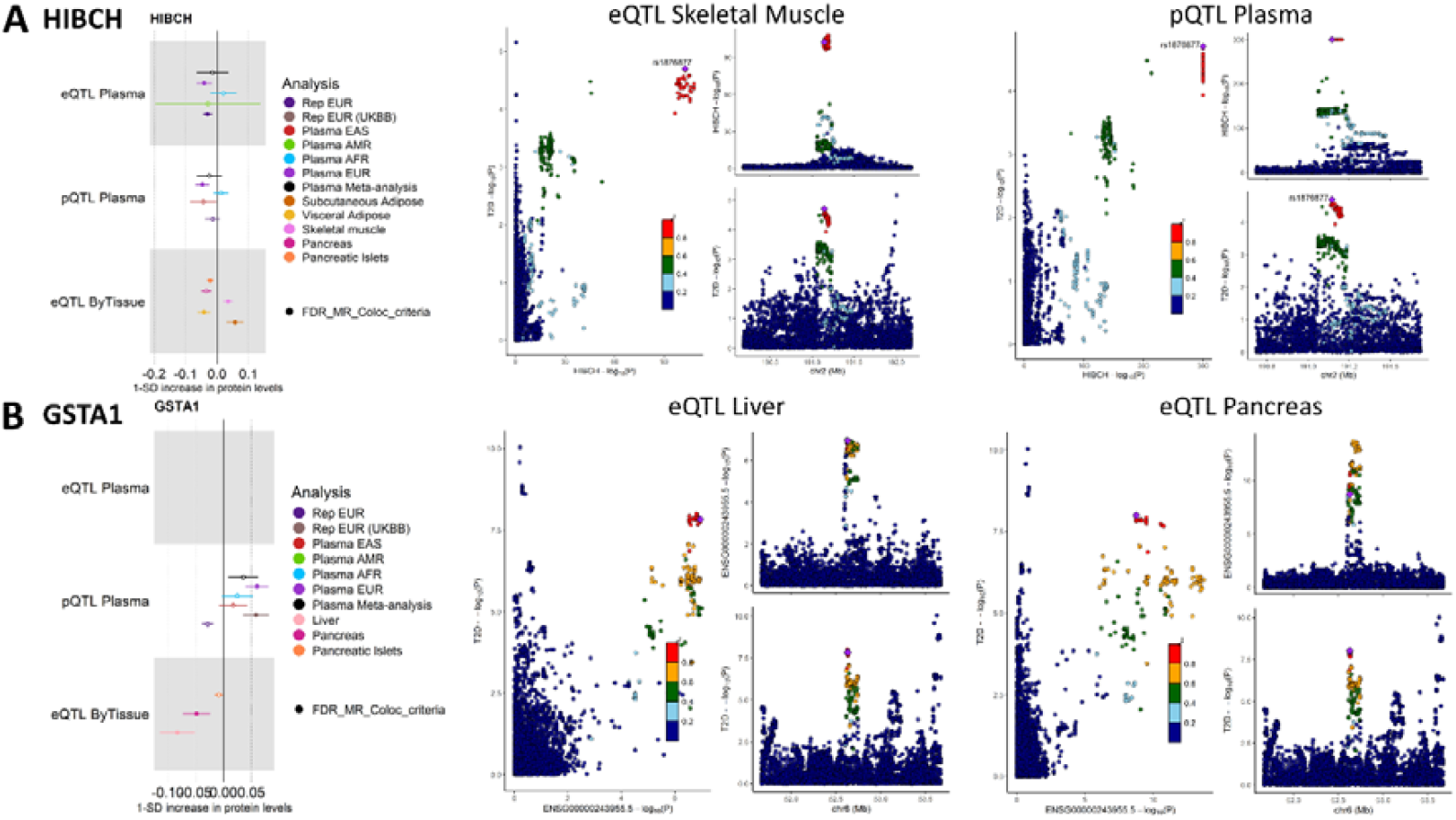
Results for HIBCH (A) and GSTA1 (B). For each molecular trait, the causal estimates from the blood eQTL, plasma pQTL, and T2D-relevant tissues eQTL MR analyses are represented, as well as the LocusCompare and LocusZoom plots demonstrating the colocalization evidence from eQTL in T2D-relevant tissues. We report causal estimates as odds ratios (OR) for T2D per standard deviation (SD) change in genetically predicted gene expression or protein levels.

Six additional molecular traits presented significant causal effects on T2D risk in both eQTL from T2D-relevant tissues and in the blood pQTL MR analyses. While we have investigated non-blood tissue MR in EUR only, due to data availability, we found overlap with blood pQTL findings from EAS for *FAM20B* and *BOC*. Future studies are needed to characterize non-blood molecular QTLs in non-EUR to validate these results. Obtaining non-blood QTLs in global populations will also provide insights into the ancestry-related heterogeneity in non-blood tissues.

Only for subcutaneous adipose tissue, skeletal muscle and liver, we found a significant enrichment of overlapping molecular traits with causal effects in blood eQTL and pQTL causal inference analyses (Methods). This highlights the non-redundant information captured by gene expression and protein abundance levels, for example due to post-transcriptional changes not captured at the RNA level^27^ (Methods, Supplemental Table 8).

### Identification of new putative candidate effector genes for T2D

We identify new causal associations between molecular traits and T2D due to the inclusion of diverse global populations. This is the case for *PTGES2* (Supplemental Figure 2), a prostaglandin E synthase associated with fatty acid metabolism and the innate immune system, widely expressed across tissues, and for which the causal effect was replicated in an independent cohort from the same genetic ancestry group^31^. Two nonsynonymous coding variants in this gene have been reported to be associated with a decreased risk of T2D in the German population^32^, as well as rs7872702, a variant upstream *PTGES2*, in the latest T2DGGI AFR GWAS meta-analysis^2^ (OR = 0.96, p-value = 5.97×10^7^). Additionally, a study has described an increase in PTGES2 protein levels in skeletal muscle after exercise in T2D patients from an Australian population^33^, a direction concordant with our MR estimates. Another example is FAM20B, for which decreased protein abundance was causally associated with increased T2D risk in EAS (**Figure 3B**). This protein was recently found to be altered in the matrisome of adipose tissues in obesity. The matrisome encompasses proteins of the extracellular matrix that play an important role in adipocyte differentiation and function^34^. Increased gene expression levels of *TUFM* (Supplemental Figure 5) were causally associated with increased T2D risk in the blood meta-analysis (OR=1.03, q-value=2.20×10^2^), but in none of the single-ancestry MR analyses (q-value EUR=5.10×10^2^, AFR=0.94, AMR=0.58). Causal effects of this gene on T2D risk were also found in the brain hypothalamus (OR=1.12, q-value=5.82×10^9^), whole pancreas (OR=1.06, q-value=2.74×10^9^) and visceral adipose tissue (OR=0.77, q-value=2.11×10^9^). *TUFM* has a suggested role downstream of the insulin cascade^35^ and its expression might be impacted by a genetic inversion associated with obesity^36^.

We identified risk-increasing causal effects of CPXM1 on T2D risk in the plasma pQTL MR analysis (OR=1.05, q-value=1.45×10^2^) and replicated in two independent cohorts from EUR, as well as in the eQTL MR analysis in skeletal muscle (OR=1.07, q-value=1.23×10^4^) and subcutaneous adipose tissue (OR=1.07, q-value=5.69×10^5^) (Supplemental Figure 5). *CPXM1* is a widely expressed gene, with a role in adipose tissue production^37^ and association with insulin resistance in polycystic ovary syndrome^38^. The additional insights obtained from T2D-primary tissues are also illustrated with *C1GALT1* which is causally associated with T2D risk in skeletal muscle (OR=1.04, q-value=5.29×10^3^) and pancreatic islets (OR=1.05, q-value=7.40×10^6^), corroborated by colocalization evidence only when using a method that tests for multiple shared signals in the investigated genomic region. GWAS of fat cell numbers identified a locus overlapping *C1GALT1*^39^, and a knock-out mouse model for this gene showed consequences on metabolism (http://www.informatics.jax.org/, https://www.mousephenotype.org/data/genes/).

## Discussion

In this study, we unravel causal molecular mechanisms influencing T2D risk in an ancestry- and tissue-aware manner. To our knowledge, this is the first multi-ancestry analysis looking at causal effects of genes and proteins on T2D risk, and the first one to integrate EAS pQTL data. In total, we have identified 963 molecular traits with a causal effect on T2D risk in at least one tissue. The causal effects of 79 gene and protein expression levels in blood were replicated in independent cohorts from a similar genetic ancestry group, including new potential T2D effector genes such as *CPXM1*, *PTGES1* and *FAM20B*. Overall, we observed low ancestry-related heterogeneity of causal effects, suggesting they are likely shared across genetic ancestry groups. Conversely, we observed high heterogeneity of causal effects across tissues, including opposite directions of effect. This was observed even between anatomically similar tissues such as pancreatic islets and the whole pancreas, potentially suggesting cell-type specific causal effects. Additionally, 85% of the causal effects of gene expression detected in T2D-relevant tissues were not detected in the blood. This highlights the need to continue increasing the granularity in measuring gene expression levels across tissues and at the single-cell level to understand biological processes leading to the development of diseases.

In addition to providing new insights on the molecular causes modifying T2D risk, we provide key methodological aspects to be integrated into similar studies for complex traits. This includes the importance of accounting for complex polygenic architectures of diseases using dedicated methods such as the colocalization approach PWCoCo^40^, and of using ancestry-matched summary statistics. For instance, when comparing the genes tested in our study with those tested in the most recent large-scale colocalization work based on the coloc method (same QTL datasets tested)^16^, we describe more colocalization signals for all the investigated T2D-relevant tissues (Supplemental Figure 6). The additional signals detected in our study map to loci with complex LD patterns such as *C1GALT1* (Supplemental Figure 7). Similarly, a prior pQTL MR study on T2D risk described causal effects for 11 proteins using the same pQTL deCODE dataset and similar criteria^9^. Our analytical approach replicated seven of these causal effects and identified additional causal effects for 39 proteins. Validating MR results through sensitivity analyses and colocalization is needed to avoid declaring false positive results. In our study, increasing the number of criteria to declare significant MR results led to an increased replication rate (Supplemental Table 9), in line with previous recommendations^4^. Despite employing a strict strategy to define putative causal effects, the replication rate among genes that could be tested in independent cohorts remained lower than 50%, potentially due to the violation of MR assumptions or power. We therefore advocate for careful interpretation of MR results and the use of replication approaches, which will be needed to validate the potential candidates from our non-blood MR results. However, replication is currently challenged by the lack of common instrumented molecular traits between the discovery and replication cohorts, worsened when molecular traits are measured on targeted panels such as in proteomics profiling.

In summary, we have conducted the largest multi-tissue and multi-ancestry causal inference analysis of T2D to date. We identify 923 genes and 46 proteins for which expression levels are causally associated with T2D risk. These findings expand the catalog of putative causal molecular traits and effector genes influencing T2D. By providing replication in independent cohorts and comparison of findings across ancestries, tissues, and molecular levels, we provide strong causal candidates modulating T2D risk, that likely generalize to many diverse global populations. Our findings help prioritize genes and proteins for investigation as molecular targets for T2D treatment or prevention.

## Supporting information

STROBE guidelines

## Methods

### Datasets

We performed two-sample MR using eQTL as well as pQTL datasets of various genetic ancestry groups and tissues (**Figure 1**, Supplemental Table 1). Blood eQTL data were available in EUR, AFR and AMR, while plasma pQTL data were available for EUR, AFR and EAS. Although genetic ancestry is on a continuous scale, we defined genetic ancestry groups based on 1000 Genomes Project phase 3 individuals as a reference^13^. GWAS summary statistics for T2D, the outcome in our MR analyses, correspond to the T2DGGI GWAS summary statistics^2^ from the same genetic ancestry group as the QTL datasets used as the exposure. When multiple QTL datasets were available for the same genetic ancestry group, we chose to maximize the sample size of the discovery cohorts. This led to the inclusion of eQTLGen^41^ for discovery and GENOA^42^ for replication in the eQTL EUR MR, and deCODE^43^ for discovery and ARIC^44^ for replication in the pQTL EUR MR. Additionally, we performed eQTL MR on seven further T2D-relevant tissues: subcutaneous adipose tissue, visceral adipose tissue, liver, brain hypothalamus, skeletal muscle, pancreas (all coming from GTEx^31^), and pancreatic islets (TIGER^45^). Due to data availability, these analyses were restricted to EUR, with T2DGGI GWAS summary statistics specific to EUR being used for the outcome data.

### Mendelian Randomization (MR)

#### Mendelian randomization assumptions

MR relies on three core assumptions, namely the relevance, independence and exclusion restriction assumptions. The relevance criteria assumes that the IVs selected to run the MR analyses should be strongly associated with the exposure, i.e., being strong predictors of the exposure. This assumption can be directly tested in MR as the strength of association between genetic variants and the exposure can be computed using the F-statistics defined as 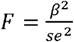, where *β* corresponds to effect size estimate of the association between the genetic variant and the gene expression or protein abundance level, and *se* to its standard deviation. The independence assumption states that the IVs are not associated with confounders of the exposure-outcome relationship. The exclusion restriction assumption ensures that there is no horizontal pleiotropy, i.e., that the effect of the IVs on the outcome are mediated solely through the exposure. Both assumptions cannot be directly tested in MR analyses, but sensitivity methods (detailed below) that relax MR assumptions can be employed to limit potential biases.

#### Selection of IVs

We selected IVs for gene expression and protein abundances from *cis*-QTL by using the cis-windows and the significance threshold defined in each study (Supplemental Table 1). To select the IVs to be used in MR, we first performed a linkage disequilibrium clumping (LD) using PLINK (version 1.9)^46^ and the default parameters recommended in the TwoSampleMR package^47^, i.e. in a 10Mb region considering an r^2^ threshold of 0.001, with the 1000 Genomes Project phase 3^13^ from the corresponding genetic ancestry group being used as the reference panel. If any IV was not present in the T2DGGI GWAS summary statistics, we replaced it with an LD-based proxy (r2 >0.8) using the *ieugwasr::ld_matrix()* R function (v1.0.2) and the output from PLINK (version 1.9)^46^ which clumps variants into clusters. We then selected variants with an F-statistic greater than 10 to limit weak instrument bias^48^ and verify the MR relevance assumption.

### Single-ancestry two-sample MR analyses

All the MR analyses were performed using the TwoSampleMR package^47^ (version 0.5.9). Alleles were harmonized between the exposure and outcome data using the function *harmonise_data()* with default parameters. We report causal estimates as OR per unit of genetically predicted gene expression and protein levels on T2D risk. We defined genes with significant causal effects as having a q-value lower than 5% from the IVW method or from the Wald ratio if only one IV was present. FDR correction was applied within each cohort. We used four sensitivity analyses: weighted median, MR-PRESSO^49^ and MR-Egger^50^ to test for pleiotropy, as well as Steiger-filtered IVW to limit for reverse causation, i.e, when the direction of the causal association is actually from the outcome to the exposure. If the distortion test of MR-PRESSO was significant (p-value < 0.05), we considered the effect estimate of the outlier-corrected method. Otherwise, we used the estimate of the raw MR-PRESSO method. We considered only molecular traits having a concordant direction of causal effect with all the sensitivity analyses, as well as showing no significant heterogeneity (heterogeneity I^2^ lower than 50%) and pleiotropy (MR-Egger intercept test p-value < 0.05). Finally, to limit the risk of false positives that could arise due to violations of the MR assumptions^19^, we performed colocalization using PWCoCo^40^, and only retained genes and proteins with a posterior probability of a share causal variant (PPH4) greater than 0.8. We note that PWCoCo first performs standard colocalization using coloc, and only when PPH4 < 0.8 does it proceed to colocalization on secondary genetic associations, found through conditional analyses. While overlap of exposure and outcome data is possible in our two-sample MR analyses, we expect the impact to be limited due to the selection of IVs highly predictive of the exposure and the high sample sizes^51^.

### Replication in independent cohorts

We tested the genes and proteins with significant causal effects in the discovery analyses for replication in independent cohort from the same genetic ancestry group (**Figure 1**, Supplemental Table 1). For the pQTL analyses, no replication cohort with the same SomaLogic platform was available in EAS. In addition to using Somalogic replication cohorts for EUR and AFR, we therefore used the UKB cohort for all populations (EUR, AFR and EAS)^21^, for which proteins were assayed using the Olink panel. We declared causal effects of genes and proteins as replicated if they showed a q-value lower than 5% (the correction being applied among the genes and proteins with significant causal effects identified in the discovery analyses and that could be tested in the replication set), and a concordant direction of effect with the IVW estimate from the discovery cohort.

### Multi-ancestry MR meta-analysis

In addition to performing single-ancestry MR, we conducted eQTL and pQTL multi-ancestry MR meta-analyses for molecular traits that could be tested in at least two ancestries using the metafor package^52^ (version 4.6). We used IVW with a random-effect model. We defined genes and proteins with significant causal effects if they presented a q-value lower than 5% in the meta-analysis, and compelling evidence in at least one cohort, i.e., with a nominal significant p-value, along with the MR sensitivity and colocalization criteria met in at least one of the entering cohorts.

### Follow-up analyses

#### Enrichment of causal effects between eQTL and pQTL MR analyses

To assess whether there was a significant enrichment of causal effects detected in both eQTL and pQTL MR analyses, we performed a Fisher’s exact test. We compared the proportion of molecular traits tested in both eQTL and pQTL MR analyses to the proportion of molecular traits with significant causal effects in both eQTL and pQTL MR analyses. This test was performed for each T2D-relevant tissue, and for blood.

#### Replication rate according to MR significance criteria

We evaluated the impact of a stricter definition of significant causal effects in our MR analyses on the replication rate by using the blood eQTL EUR data as an example, as it presents the largest number of significant findings (eQTLGen as the discovery cohort and GENOA as the replication cohort). We computed the replication rate by defining significant signals in the discovery cohort using 5 criteria: **(1)** all tested genes as significant, **(2)** genes with a p-value nominally significant, **(3)** genes with a q-value lower than 5%, **(4)** genes from **(3)** with a concordant direction of effect with sensitivity analyses, no heterogeneity and no pleiotropy **(5)** genes from **(4)** with evidence from colocalization (PPH4>0.8). For the latter, we recomputed the replication rate by distinguishing genes with only one IV from genes with more than one IV given that no sensitivity methods can be applied if only one IV is present. Replication was tested for the significant causal effects in the discovery cohort, and the replication rate was computed as the proportion of genes with a q-value lower than 5% in the replication cohort among the tested genes for each replication criteria. The FDR-correction in the replication cohort was applied to the set of significant genes from the discovery cohort that could be tested in the replication cohort.

### Comparisons with findings from the latest large-scale T2D colocalization study

We compared our findings to the results from the latest large-scale colocalization effort^16^ which used the T2DGGI GWAS multi-ancestry summary statistics^2^ and the coloc method^17^. The authors employed an approach to test for colocalization around the 1,289 T2D index variants. Our approach was centered on the molecular traits, as we investigated causal effects through MR corroborated by colocalization for all the molecular traits that could be instrumented. We restricted our comparisons to the molecular traits tested in both studies. We only compared the analyses in T2D-relevant tissues across the two studies for the subcutaneous adipose tissue, visceral adipose tissue, liver, brain hypothalamus, skeletal muscle and pancreatic islets as they were based on the same cohorts (GTEx and TIGER).

## Author contributions

O.B, A.L.A, S.Y, A.P.M and E.Z conceived the project and designed analysis plan which was reviewed by all co-authors. S.Y, T-Y. Y and F.M provided pQTL data in EAS. O.B, A.L.A and S.Y developped the analysis pipeline. O.B, A.L.A, S.Y and C.Z performed the analyses. O.B and S.Y created the figures. O.B and A.L.A drafted the manuscript, with inputs from all co-authors.

## Acknowledgments

OB has received funding from the European Union’s Horizon 2020 research and innovation programme under Grant Agreement No 101017802 (OPTOMICS). SY has received funding from the Japan Society for the Promotion of Science (202460267) and Canadian Institutes of Health Research (AD6-200177). This work is supported in part by the National Center for Advancing Translational Sciences, CTSI grant UL1TR001881, and the National Institute of Diabetes and Digestive and Kidney Disease Diabetes Research Center (DRC) grant DK063491 to the Southern California Diabetes Endocrinology Research Center. Infrastructure for the CHARGE Consortium is supported in part by the National Heart, Lung, and Blood Institute (NHLBI) grant R01HL105756. The TIGER data have been generated by the T2DSystems Consortium, and downloaded from the TIGER Data Portal - http://tiger.bsc.es. CNS was supported by the National Institutes of Health (R01DK118011 and R01DK136671) and the American Diabetes Association (11-22-JDFPM-06).

## Conflict of interest

The authors declare no conflict of interest.

## Data availability

All contributing cohorts have ethical approval from their institutional ethics review boards. All data used in the study, except for the Kyoto cohort (EAS pQTL), are publicly available with reference to the corresponding studies summarized in Supplemental Table 1. The Kyoto Nagahama cohort study data will be available after publication of the study at https://www.hgvd.genome.med.kyoto-u.ac.jp/.

## Code availability

The code used to perform all the single-ancestry MR analyses and the MR meta-analysis across genetic ancestry groups are available at https://github.com/Ozvan/OmicsMR.

## Supplemental Tables

**Supplemental Table 1:** Summary of the QTL data used to select IVs for the MR analyses, including genetic ancestry group, sample size, tissue, corresponding publication and cis-window definition. TSS: Transcription Start Site, FDR: False Discovery Rate.

**Supplemental Table 2:** Results of the blood eQTL MR analysis in the meta-analysis across ancestries and in all genetic ancestry groups assessed (EUR, AFR, AMR). For the meta-analysis results, the number of genetic ancestry groups entering the meta-analysis and the I^2^ value are provided. For the single-ancestry MR analyses, the number of IVs, the MR method used and the PPH4 value from PWCoCo are indicated.

**Supplemental Table 3:** Results of the blood pQTL MR analysis in the meta-analysis across ancestries and in all genetic ancestry groups assessed (EUR, AFR, EAS). For the meta-analysis results, the number of genetic ancestry groups entering the meta-analysis and the I^2^ value are provided. For the single-ancestry MR analyses, the number of IVs, the MR method used and the PPH4 value from PWCoCo are indicated.

**Supplemental Table 4:** Results of the blood eQTL MR analysis in the replication cohorts. Only the genes with significant causal effects in the discovery cohort are reported. For comparison purposes, the results from the blood eQTL disvoery MR analyses in the corresponding genetic ancestry group are also reported.

**Supplemental Table 5:** Results of the blood pQTL MR analysis in the replication cohorts. Only the proteins with significant causal effects in the discovery cohort are reported. For comparison purposes, the results from the blood pQTL disvoery MR analyses in the corresponding genetic ancestry group are also reported.

**Supplemental Table 6:** Number of genes and proteins with significant causal effects in the discovery MR analyses in each genetic ancestry group. The number of gene and proteins from this set tested for replication and with replicated causal effects are also reported.

**Supplemental Table 7:** Results of the eQTL MR analysis in the seven T2D-relevant tissues (only in EUR due to data availability). The number of IVs, the MR method used and the PPH4 value from PWCoCo are indicated.

**Supplemental Table 8:** Chi-square test comparing the overlap between genes with significant causal effects from the eQTL and pQTL MR analyses among the tested proteins and the proteins with significant causal effects. The test has been performed for each tissue.

**Supplemental Table 9:** Number of genes with significant causal effects using different criteria detected in the EUR discovery cohort (eQTLGen) and tested and replicated in the EUR replication cohort (GENOA). The final column corresponds to the criteria employed in our study, and distinguishes the genes with only one or more than one IVs. The causal effect of a gene is considered as replicated if it has a q-value lower than 5% among the genes tested for replication with a consistent direction of effect with the discovery analysis.

## Supplemental Figures

**Supplemental Figure 1:** Forest plots of the causal effects identified in the blood eQTL MR analyses for genes detected only in the meta-analysis but not in single-ancestry MR analyses. Causal estimates from the single-ancestry MR in the discovery cohorts (matched genetic ancestry group between the exposure and the outcome) are also represented. Filled dots represent causal estimates from MR analyses that have a q-value<0.05, and (1) pass the sensitivity criteria and show evidence of colocalization (PPH4>0.8) in single-ancestry analyses, or (2) present nominal significance and meet criteria (1) in at least one cohort entering the meta-analysis. Genes and proteins with causal effects identified in single-ancestry analyses and replicated in independent cohorts from the same genetic ancestry group are denoted with a star. We report causal estimates as odds ratios (OR) for T2D per standard deviation (SD) change in genetically predicted gene expression or protein levels.

**Supplemental Figure 2:** Causal estimates and frequency and effect sizes of IVs on *TOLLIP-AS1* and *PTGES2* (A) Forest plots of causal estimates for *TOLLIP-AS1* and *PTGES2* from the eQTL MR analyses. Causal estimates from the single-ancestry MR in the discovery cohorts (matched genetic ancestry group between the exposure and the outcome) are also represented. Filled dots represent causal estimates from MR analyses that have a q-value<0.05, and (1) pass the sensitivity criteria and show evidence of colocalization (PPH4>0.8) in single-ancestry analyses, or (2) present nominal significance and meet criteria (1) in at least one cohort entering the meta-analysis. Genes and proteins with causal effects identified in single-ancestry analyses and replicated in independent cohorts from the same genetic ancestry group are denoted with a star. We report causal estimates as odds ratios (OR) for T2D per standard deviation (SD) change in genetically predicted gene expression or protein levels. (B) LocusCompare and LocusZoom for the two genes demonstrating colocalization evidence. (C) Allele frequency of the IVs selected for the two genes, and their association with the corresponding gene expression levels.

**Supplemental Figure 3:** LocusCompare and LocusZoom for FAM20B and BOC, two proteins with significant causal effects detected only in EAS. Results are displayed in EUR (top panel) and in EAS (bottom panel).

**Supplemental Figure 4:** Distribution of significant causal effect estimates according to the tissue of the eQTL dataset.

**Supplemental Figure 5:** Forest plots of causal estimates for TUFM and CPXM1 from all the MR analyses. Causal estimates from the single-ancestry MR in the discovery cohorts (matched genetic ancestry group between the exposure and the outcome) are also represented. Filled dots represent causal estimates from MR analyses that have a q-value<0.05, and (1) pass the sensitivity criteria and show evidence of colocalization (PPH4>0.8) in single-ancestry analyses, or (2) present nominal significance and meet criteria (1) in at least one cohort entering the meta-analysis. Genes and proteins with causal effects identified in single-ancestry analyses and replicated in independent cohorts from the same genetic ancestry group are denoted with a star. We report causal estimates as odds ratios (OR) for T2D per standard deviation (SD) change in genetically predicted gene expression or protein levels.

**Supplemental Figure 6:** Venn Diagrams representing the overlap of genes identified in the previous colocalization study from Mandla et al. 2024, and in the present study (MR+colocalization using PWCoCo). Comparisons are indicated for six tissues which used the same QTL datasets in both studies.

**Supplemental Figure 7:** LocusCompare and LocusZoom for *C1GALT1* in Skeletal Muscle and in Pancreatic Islets. PPH4 obtained with the coloc approach (from Mandla et al. 2024) and with the PWCoCo approach (the present study) are indicated.

## Notes

### Competing Interest Statement

The authors have declared no competing interest.

### Funding Statement

OB has received funding from the European Union Horizon 2020 research and innovation programme under Grant Agreement No 101017802 (OPTOMICS). SY has received funding from the Japan Society for the Promotion of Science (202460267) and Canadian Institutes of Health Research (AD6-200177). This work is supported in part by the National Center for Advancing Translational Sciences, CTSI grant UL1TR001881, and the National Institute of Diabetes and Digestive and Kidney Disease Diabetes Research Center (DRC) grant DK063491 to the Southern California Diabetes Endocrinology Research Center. Infrastructure for the CHARGE Consortium is supported in part by the National Heart, Lung, and Blood Institute (NHLBI) grant R01HL105756. The TIGER data have been generated by the T2DSystems Consortium, and downloaded from the TIGER Data Portal - http://tiger.bsc.es. CNS was supported by the National Institutes of Health (R01DK118011 and R01DK136671) and the American Diabetes Association (11-22-JDFPM-06).

### Author Declarations

All contributing cohorts have ethical approval from their institutional ethics review boards. All data used in the study, except for the Kyoto cohort (EAS pQTL), are publicly available with reference to the corresponding studies summarized in Supplemental Table 1. The Kyoto Nagahama cohort study was approved by the ethics committees of Kyoto University Graduate School of Medicine and the Nagahama Municipal Review Board (No. 278). The Kyoto Nagahama cohort study data will be available after publication of the study at https://www.hgvd.genome.med.kyoto-u.ac.jp/.

## References

1 Mahajan, A. et al. Multi-ancestry genetic study of type 2 diabetes highlights the power of diverse populations for discovery and translation. Nat Genet 54, 560–572 (2022). 10.1038/s41588-022-01058-3

2 Suzuki, K. et al. Genetic drivers of heterogeneity in type 2 diabetes pathophysiology. Nature (2024). 10.1038/s41586-024-07019-6

3 Pearson, E. R. Type 2 diabetes: a multifaceted disease. Diabetologia 62, 1107–1112 (2019). 10.1007/s00125-019-4909-y

4 Zhao, H. et al. Proteome-wide Mendelian randomization in global biobank meta-analysis reveals multi-ancestry drug targets for common diseases. Cell Genom 2, None (2022). 10.1016/j.xgen.2022.100195

5 Davey Smith, G. & Hemani, G. Mendelian randomization: genetic anchors for causal inference in epidemiological studies. Hum Mol Genet 23, R89–98 (2014). 10.1093/hmg/ddu328

6 Yousri, N. A., Albagha, O. M. E. & Hunt, S. C. Integrated epigenome, whole genome sequence and metabolome analyses identify novel multi-omics pathways in type 2 diabetes: a Middle Eastern study. BMC Med 21, 347 (2023). 10.1186/s12916-023-03027-x

7 Chung, R.-H. et al. Elucidating the Epigenetic Landscape of Type 2 Diabetes: A Multi-Omics Analysis Revealing Novel CpG Sites and Their Association with Cardiometabolic Traits. medRxiv, 2024.2005.2020.24307650 (2024). 10.1101/2024.05.20.24307650

8 de Klerk, J. A. et al. Altered blood gene expression in the obesity-related type 2 diabetes cluster may be causally involved in lipid metabolism: a Mendelian randomisation study. Diabetologia 66, 1057–1070 (2023). 10.1007/s00125-023-05886-8

9 Yuan, S. et al. Plasma proteins and onset of type 2 diabetes and diabetic complications: Proteome-wide Mendelian randomization and colocalization analyses. Cell Rep Med 4, 101174 (2023). 10.1016/j.xcrm.2023.101174

10 Porcu, E. et al. Triangulating evidence from longitudinal and Mendelian randomization studies of metabolomic biomarkers for type 2 diabetes. Sci Rep 11, 6197 (2021). 10.1038/s41598-021-85684-7

11 Bocher, O. et al. Disentangling the consequences of type 2 diabetes on targeted metabolite profiles using causal inference and interaction QTL analyses. PLoS Genet 20, e1011346 (2024). 10.1371/journal.pgen.1011346

12 Kreitmaier, P., Katsoula, G. & Zeggini, E. Insights from multi-omics integration in complex disease primary tissues. Trends Genet 39, 46–58 (2023). 10.1016/j.tig.2022.08.005

13 Genomes Project, C. et al. A global reference for human genetic variation. Nature 526, 68–74 (2015). 10.1038/nature15393

14 Pozarickij, A. et al. Causal relevance of different blood pressure traits on risk of cardiovascular diseases: GWAS and Mendelian randomisation in 100,000 Chinese adults. Nat Commun 15, 6265 (2024). 10.1038/s41467-024-50297-x

15 Ken, S. et al. Multi-ancestry genome-wide study in >2.5 million individuals reveals heterogeneity in mechanistic pathways of type 2 diabetes and complications. medRxiv, 2023.2003.2031.23287839 (2023). 10.1101/2023.03.31.23287839

16 Mandla, R. et al. Multi-omics characterization of type 2 diabetes associated genetic variation. medRxiv, 2024.2007.2015.24310282 (2024). 10.1101/2024.07.15.24310282

17 Giambartolomei, C. et al. Bayesian test for colocalisation between pairs of genetic association studies using summary statistics. PLoS Genet 10, e1004383 (2014). 10.1371/journal.pgen.1004383

18 Gkatzionis, A., Burgess, S. & Newcombe, P. J. Statistical methods for cis-Mendelian randomization with two-sample summary-level data. Genet Epidemiol 47, 3–25 (2023). 10.1002/gepi.22506

19 Zuber, V. et al. Combining evidence from Mendelian randomization and colocalization: Review and comparison of approaches. Am J Hum Genet 109, 767–782 (2022). 10.1016/j.ajhg.2022.04.001

20 Arruda, A. L., Morris, A. P. & Zeggini, E. Advancing equity in human genomics through tissue-specific multi-ancestry molecular data. Cell Genom 4, 100485 (2024). 10.1016/j.xgen.2023.100485

21 Sudlow, C. et al. UK biobank: an open access resource for identifying the causes of a wide range of complex diseases of middle and old age. PLoS Med 12, e1001779 (2015). 10.1371/journal.pmed.1001779

22 Hu, S. et al. Leveraging fine-scale population structure reveals conservation in genetic effect sizes between human populations across a range of human phenotypes. bioRxiv, 2023.2008.2008.552281 (2023). 10.1101/2023.08.08.552281

23 Costanzo, M. C. et al. The Type 2 Diabetes Knowledge Portal: An open access genetic resource dedicated to type 2 diabetes and related traits. Cell Metab 35, 695–710 e696 (2023). 10.1016/j.cmet.2023.03.001

24 Rottner, A. K. et al. A genome-wide CRISPR screen identifies CALCOCO2 as a regulator of beta cell function influencing type 2 diabetes risk. Nat Genet 55, 54–65 (2023). 10.1038/s41588-022-01261-2

25 Karczewski, K. J. et al. The mutational constraint spectrum quantified from variation in 141,456 humans. Nature 581, 434–443 (2020). 10.1038/s41586-020-2308-7

26 Consortium, G. T. et al. Genetic effects on gene expression across human tissues. Nature 550, 204–213 (2017). 10.1038/nature24277

27 Buccitelli, C. & Selbach, M. mRNAs, proteins and the emerging principles of gene expression control. Nat Rev Genet 21, 630–644 (2020). 10.1038/s41576-020-0258-4

28 Gudmundsdottir, V. et al. Circulating Protein Signatures and Causal Candidates for Type 2 Diabetes. Diabetes 69, 1843–1853 (2020). 10.2337/db19-1070

29 Li, Y. et al. Mitochondrial dysfunction and onset of type 2 diabetes along with its complications: a multi-omics Mendelian randomization and colocalization study. Front Endocrinol (Lausanne) 15, 1401531 (2024). 10.3389/fendo.2024.1401531

30 Rooney, M. R. et al. Proteomic Predictors of Incident Diabetes: Results From the Atherosclerosis Risk in Communities (ARIC) Study. Diabetes Care 46, 733–741 (2023). 10.2337/dc22-1830

31 Consortium, G. T. The Genotype-Tissue Expression (GTEx) project. Nat Genet 45, 580–585 (2013). 10.1038/ng.2653

32 Nitz, I. et al. Association of prostaglandin E synthase 2 (PTGES2) Arg298His polymorphism with type 2 diabetes in two German study populations. J Clin Endocrinol Metab 92, 3183–3188 (2007). 10.1210/jc.2006-2550

33 Hussey, S. E. et al. Effect of exercise on the skeletal muscle proteome in patients with type 2 diabetes. Med Sci Sports Exerc 45, 1069–1076 (2013). 10.1249/MSS.0b013e3182814917

34 Kaartinen, M. T. et al. Matrisome alterations in obesity - Adipose tissue transcriptome study on monozygotic weight-discordant twins. Matrix Biol 108, 1–19 (2022). 10.1016/j.matbio.2022.02.005

35 Mercader, J. M. et al. Identification of novel type 2 diabetes candidate genes involved in the crosstalk between the mitochondrial and the insulin signaling systems. PLoS Genet 8, e1003046 (2012). 10.1371/journal.pgen.1003046

36 Gonzalez, J. R. et al. Polymorphic Inversions Underlie the Shared Genetic Susceptibility of Obesity-Related Diseases. Am J Hum Genet 106, 846–858 (2020). 10.1016/j.ajhg.2020.04.017

37 Kim, Y. H. et al. Identification of carboxypeptidase X (CPX)-1 as a positive regulator of adipogenesis. FASEB J 30, 2528–2540 (2016). 10.1096/fj.201500107R

38 Pervaz, S. et al. Role of CPXM1 in Impaired Glucose Metabolism and Ovarian Dysfunction in Polycystic Ovary Syndrome. Reprod Sci 30, 526–543 (2023). 10.1007/s43032-022-00987-y

39 Kulyte, A., Aman, A., Strawbridge, R. J., Arner, P. & Dahlman, I. A. Genome-Wide Association Study Identifies Genetic Loci Associated With Fat Cell Number and Overlap With Genetic Risk Loci for Type 2 Diabetes. Diabetes 71, 1350–1362 (2022). 10.2337/db21-0804

40 Zheng, J. et al. Phenome-wide Mendelian randomization mapping the influence of the plasma proteome on complex diseases. Nat Genet 52, 1122–1131 (2020). 10.1038/s41588-020-0682-6

41 Vosa, U. et al. Large-scale cis- and trans-eQTL analyses identify thousands of genetic loci and polygenic scores that regulate blood gene expression. Nat Genet 53, 1300–1310 (2021). 10.1038/s41588-021-00913-z

42 Shang, L. et al. Genetic Architecture of Gene Expression in European and African Americans: An eQTL Mapping Study in GENOA. Am J Hum Genet 106, 496–512 (2020). 10.1016/j.ajhg.2020.03.002

43 Ferkingstad, E. et al. Large-scale integration of the plasma proteome with genetics and disease. Nat Genet 53, 1712–1721 (2021). 10.1038/s41588-021-00978-w

44 Zhang, J. et al. Plasma proteome analyses in individuals of European and African ancestry identify cis-pQTLs and models for proteome-wide association studies. Nat Genet 54, 593–602 (2022). 10.1038/s41588-022-01051-w

45 Alonso, L. et al. TIGER: The gene expression regulatory variation landscape of human pancreatic islets. Cell Rep 37, 109807 (2021). 10.1016/j.celrep.2021.109807

46 Purcell, S. et al. PLINK: a tool set for whole-genome association and population-based linkage analyses. Am J Hum Genet 81, 559–575 (2007). 10.1086/519795

47 Hemani, G. et al. The MR-Base platform supports systematic causal inference across the human phenome. Elife 7 (2018). 10.7554/eLife.34408

48 Lawlor, D. A., Harbord, R. M., Sterne, J. A., Timpson, N. & Davey Smith, G. Mendelian randomization: using genes as instruments for making causal inferences in epidemiology. Stat Med 27, 1133–1163 (2008). 10.1002/sim.3034

49 Verbanck, M., Chen, C. Y., Neale, B. & Do, R. Detection of widespread horizontal pleiotropy in causal relationships inferred from Mendelian randomization between complex traits and diseases. Nat Genet 50, 693–698 (2018). 10.1038/s41588-018-0099-7

50 Bowden, J., Davey Smith, G. & Burgess, S. Mendelian randomization with invalid instruments: effect estimation and bias detection through Egger regression. Int J Epidemiol 44, 512–525 (2015). 10.1093/ije/dyv080

51 Mounier, N. & Kutalik, Z. Bias correction for inverse variance weighting Mendelian randomization. Genet Epidemiol 47, 314–331 (2023). 10.1002/gepi.22522

52 Viechtbauer, W. Conducting Meta-Analyses in R with the metafor Package. Journal of Statistical Software 36, 1–48 (2010). 10.18637/jss.v036.i03

