## Supplementary material for "Unravelling the molecular mechanisms causal to type 2 diabetes across global populations and disease-relevant tissues": STROBE guidelines

**STROBE-MR checklist of recommended items to address in reports of Mendelian randomization studies**

| **Item No.** | **Section** | **Checklist item** | **Page No.** | **Relevant text from manuscript** |
| --- | --- | --- | --- | --- |
| 1 | **TITLE and ABSTRACT** | Indicate Mendelian randomization (MR) as the study’s design in the title and/or the abstract if that is a main purpose of the study |  | We specified the method used for causal inference in the abstract section: “Using two-sample Mendelian randomization corroborated by colocalization” |
|  | **INTRODUCTION** |  |  |  |
| 2 | **Background** | Explain the scientific background and rationale for the reported study. What is the exposure? Is a potential causal relationship between exposure and outcome plausible? Justify why MR is a helpful method to address the study question |  | In the introduction section: ‘Here, we study the causal links of gene expression and protein abundance levels with T2D risk using MR approaches based on cis-QTLs, predicted to have the strongest biological impact on molecular traits. We expand previous causal inference studies by (1) leveraging the T2D genetic associations reported by T2DGGI and QTL maps from global populations in single-ancestry analyses and multi-ancestry meta-analyses and (2) investigating causal effects in seven further tissues relevant to T2D.’ |
| 3 | **Objectives** | State specific objectives clearly, including pre-specified causal hypotheses (if any). State that MR is a method that, under specific assumptions, intends to estimate causal effects |  | Introduction section: above text and ‘While causal inference is challenging, statistical inference methods such as Mendelian randomization (MR) can provide estimates of the causal effect of an exposure on an outcome under certain assumptions through the use of genetic instrumental variables (IVs), i.e., genetic variants predictive of the exposure.’ |
|  | **METHODS** |  |  |  |
| 4 | **Study design and data sources** | Present key elements of the study design early in the article. Consider including a table listing sources of data for all phases of the study. For each data source contributing to the analysis, describe the following: |  |  |
|  | a) | Setting: Describe the study design and the underlying population, if possible. Describe the setting, locations, and relevant dates, including periods of recruitment, exposure, follow-up, and data collection, when available. |  | In the results section, the paragraph ‘Study design overview’ provides the study design, populations under study and criteria to define statistical significance. |
|  | b) | Participants: Give the eligibility criteria, and the sources and methods of selection of participants. Report the sample size, and whether any power or sample size calculations were carried out prior to the main analysis |  | Supplemental Table 1 |
|  | c) | Describe measurement, quality control and selection of genetic variants |  | The genetic variants used were obtained from publicly available summary statistics. Selection of the IVs is presented in the paragraph ‘Selection of IVs’. |
|  | d) | For each exposure, outcome, and other relevant variables, describe methods of assessment and diagnostic criteria for diseases |  | Not relevant, publicly available summary statistics were used. |
|  | e) | Provide details of ethics committee approval and participant informed consent, if relevant |  | In the data availability section: ‘All contributing cohorts obtained ethical approval from their institutional ethics review boards.’ |
| 5 | **Assumptions** | Explicitly state the three core IV assumptions for the main analysis (relevance, independence and exclusion restriction) as well assumptions for any additional or sensitivity analysis |  | Paragraph ‘Mendelian randomization assumptions’ and ‘Single-ancestry two-sample MR analyses’ |
| 6 | **Statistical methods: main analysis** | Describe statistical methods and statistics used |  |  |
|  | a) | Describe how quantitative variables were handled in the analyses (i.e., scale, units, model) |  | The exposures are in a continuous scale was the outcome is a binary phenotype.  ‘We report causal estimates as odds ratios (OR) for T2D per standard deviation (SD) change in genetically predicted gene expression or protein levels.’ (‘Study design overview’ in the results and figure legends). |
|  | b) | Describe how genetic variants were handled in the analyses and, if applicable, how their weights were selected |  | Paragraph ‘Selection of IVs’ in the methods. |
|  | c) | Describe the MR estimator (e.g. two-stage least squares, Wald ratio) and related statistics. Detail the included covariates and, in case of two-sample MR, whether the same covariate set was used for adjustment in the two samples |  | Paragraphs ‘Single-ancestry two-sample MR analyses’ in the methods and ‘Study design overview’ in the results. |
|  | d) | Explain how missing data were addressed |  | Not applicable |
|  | e) | If applicable, indicate how multiple testing was addressed |  | FDR correction detailed in paragraphs ‘Single-ancestry two-sample MR analyses’ and ‘Multi-ancestry MR meta-analysis’ in the methods and ‘Study design overview’ in the results. |
| 7 | **Assessment of assumptions** | Describe any methods or prior knowledge used to assess the assumptions or justify their validity |  | Paragraphs ‘Mendelian randomization assumptions’, ‘Selection of IVs’ and ‘Single-ancestry two-sample MR analyses’ in the methods |
| 8 | **Sensitivity analyses and additional analyses** | Describe any sensitivity analyses or additional analyses performed (e.g. comparison of effect estimates from different approaches, independent replication, bias analytic techniques, validation of instruments, simulations) |  | Sensitivity analyses: paragraph and ‘Single-ancestry two-sample MR analyses’ in the methods.  Replication in independent cohorts: ‘Replication in independent cohorts’ in the methods.  Overall overview in ‘Study design overview’ in the results. |
| 9 | **Software and pre-registration** |  |  |  |
|  | a) | Name statistical software and package(s), including version and settings used |  | Whole method section ‘Mendelian Randomization (MR)’ |
|  | b) | State whether the study protocol and details were pre-registered (as well as when and where) |  | Not relevant |
|  | **RESULTS** |  |  |  |
| 10 | **Descriptive data** |  |  |  |
|  | a) | Report the numbers of individuals at each stage of included studies and reasons for exclusion. Consider use of a flow diagram |  | Use of summary statistics from cohorts detailed in Supplementary Table 1. We used publicly available GWAS and hence did not exclude any individual data. |
|  | b) | Report summary statistics for phenotypic exposure(s), outcome(s), and other relevant variables (e.g. means, SDs, proportions) |  | Data availability section, Supplementary Table 1 |
|  | c) | If the data sources include meta-analyses of previous studies, provide the assessments of heterogeneity across these studies |  |  |
|  | d) | For two-sample MR:  i.  Provide justification of the similarity of the genetic variant-exposure associations between the exposure and outcome samples  ii.  Provide information on the number of individuals who overlap between the exposure and outcome studies |  | The MR analyses were performed within genetic ancestry groups, and the MR estimates were meta-analysed across genetic ancestry groups (whole method section ‘Mendelian Randomization (MR)’ and ‘Study design overview’ in the results.)  Overlap between the exposure and outcome data: ‘While overlap of exposure and outcome data is possible in our two-sample MR analyses, we expect the impact to be limited due to the selection of IVs highly predictive of the exposure and the high sample sizes.’ In the methods |
| 11 | **Main results** |  |  |  |
|  | a) | Report the associations between genetic variant and exposure, and between genetic variant and outcome, preferably on an interpretable scale |  | This can be extracted from the respective publicly available GWAS outlined in the Supplementary Table 1. |
|  | b) | Report MR estimates of the relationship between exposure and outcome, and the measures of uncertainty from the MR analysis, on an interpretable scale, such as odds ratio or relative risk per SD difference |  | Results section, main figures 2, 3 and 6, supplementary figures 1, 2, 5, supplementary tables 2-5, 7.  ‘We report causal estimates as odds ratios (OR) for T2D per standard deviation (SD) change in genetically predicted gene expression or protein levels.’ (‘Study design overview’ in the results). |
|  | c) | If relevant, consider translating estimates of relative risk into absolute risk for a meaningful time period |  | Not relevant |
|  | d) | Consider plots to visualize results (e.g. forest plot, scatterplot of associations between genetic variants and outcome versus between genetic variants and exposure) |  | Main figures 2, 3 and 6  Supplementary figures 1, 2, 5, supplementary tables 2-5, 7. |
| 12 | **Assessment of assumptions** |  |  |  |
|  | a) | Report the assessment of the validity of the assumptions |  | Paragraphs ‘Mendelian randomization assumptions’, ‘Selection of IVs’ and ‘Single-ancestry two-sample MR analyses’ in the methods. |
|  | b) | Report any additional statistics (e.g., assessments of heterogeneity across genetic variants, such as *I^2^*, Q statistic or E-value) |  | Paragraphs ‘Selection of IVs’ and ‘Single-ancestry two-sample MR analyses’ in the methods  Supplementary Tables 2-3, 7 |
| 13 | **Sensitivity analyses and additional analyses** |  |  |  |
|  | a) | Report any sensitivity analyses to assess the robustness of the main results to violations of the assumptions |  | Supplementary Tables 2-3, 7  Forest plots in main figures 2, 3 and 6 and supplementary figures 1, 2, 5 |
|  | b) | Report results from other sensitivity analyses or additional analyses |  | Colocalization evidence in all results (Supplementary Tables 2-3, 7)  Replication in independent cohort (Supplementary Tables 4,5) |
|  | c) | Report any assessment of direction of causal relationship (e.g., bidirectional MR) |  | Steiger filtering as sensitivity analysis (‘Single-ancestry two-sample MR analyses’ in the methods) |
|  | d) | When relevant, report and compare with estimates from non-MR analyses |  | Paragraph ‘Identification of new putative candidate effector genes for T2D’ in the results. |
|  | e) | Consider additional plots to visualize results (e.g., leave-one-out analyses) |  |  |
|  | **DISCUSSION** |  |  |  |
| 14 | **Key results** | Summarize key results with reference to study objectives |  | Discussion section: 1^st^ paragraph |
| 15 | **Limitations** | Discuss limitations of the study, taking into account the validity of the IV assumptions, other sources of potential bias, and imprecision. Discuss both direction and magnitude of any potential bias and any efforts to address them |  | Discussion section: 2^nd^ paragraph  Also incorporated in the results section |
| 16 | **Interpretation** |  |  |  |
|  | a) | Meaning: Give a cautious overall interpretation of results in the context of their limitations and in comparison with other studies |  | Discussion section and throughout the results |
|  | b) | Mechanism: Discuss underlying biological mechanisms that could drive a potential causal relationship between the investigated exposure and the outcome, and whether the gene-environment equivalence assumption is reasonable. Use causal language carefully, clarifying that IV estimates may provide causal effects only under certain assumptions |  | Examples of specific biological mechanisms are given throughout the results and especially in the section ‘Identification of new putative candidate effector genes for T2D’.  Discussion section: ‘Despite employing a strict strategy to define putative causal effects, the replication rate among genes that could be tested in independent cohorts remained lower than 50%, potentially due to the violation of MR assumptions or power. We therefore advocate for careful interpretation of MR results and the use of replication approaches, which will be needed to validate the potential candidates from our non-blood MR results.’ |
|  | c) | Clinical relevance: Discuss whether the results have clinical or public policy relevance, and to what extent they inform effect sizes of possible interventions |  | Discussion section: ‘In summary, we have conducted the largest multi-tissue and multi-ancestry causal inference analysis of T2D to date. We identify 923 genes and 46 proteins for which expression levels are causally associated with T2D risk. These findings expand the catalog of putative causal molecular traits and effector genes influencing T2D. By providing replication in independent cohorts and comparison of findings across ancestries, tissues, and molecular levels, we provide strong causal candidates modulating T2D risk, that likely generalize to many diverse global populations. Our findings help prioritize genes and proteins for investigation as molecular targets for T2D treatment or prevention.’ |
| 17 | **Generalizability** | Discuss the generalizability of the study results (a) to other populations, (b) across other exposure periods/timings, and (c) across other levels of exposure |  | The multi-ancestry aspect is a key message of the paper, with assessment of the heterogeneity of MR estimates across genetic ancestry groups. Paragraphs ‘Low ancestry-related heterogeneity for blood molecular traits causal to T2D risk’ and ‘Assessing QTLs in global populations improves the detection of causal effects‘ and in the Discussion |
|  | **OTHER INFORMATION** |  |  |  |
| 18 | **Funding** | Describe sources of funding and the role of funders in the present study and, if applicable, sources of funding for the databases and original study or studies on which the present study is based |  | Acknowledgments |
| 19 | **Data and data sharing** | Provide the data used to perform all analyses or report where and how the data can be accessed, and reference these sources in the article. Provide the statistical code needed to reproduce the results in the article, or report whether the code is publicly accessible and if so, where |  | Data availability section  Code availability section  Supplementary Table 1 |
| 20 | **Conflicts of Interest** | All authors should declare all potential conflicts of interest |  | Section ‘conflict of interest’ |

This checklist is copyrighted by the Equator Network under the Creative Commons Attribution 3.0 Unported (CC BY 3.0) license.

1.	Skrivankova VW, Richmond RC, Woolf BAR, Yarmolinsky J, Davies NM, Swanson SA, et al. Strengthening the Reporting of Observational Studies in Epidemiology using Mendelian Randomization (STROBE-MR) Statement. JAMA. 2021;under review.

2.	Skrivankova VW, Richmond RC, Woolf BAR, Davies NM, Swanson SA, VanderWeele TJ, et al. Strengthening the Reporting of Observational Studies in Epidemiology using Mendelian Randomisation (STROBE-MR): Explanation and Elaboration. BMJ. 2021;375:n2233.
