## Supplemental Figures for "Unravelling the molecular mechanisms causal to type 2 diabetes across global populations and disease-relevant tissues"

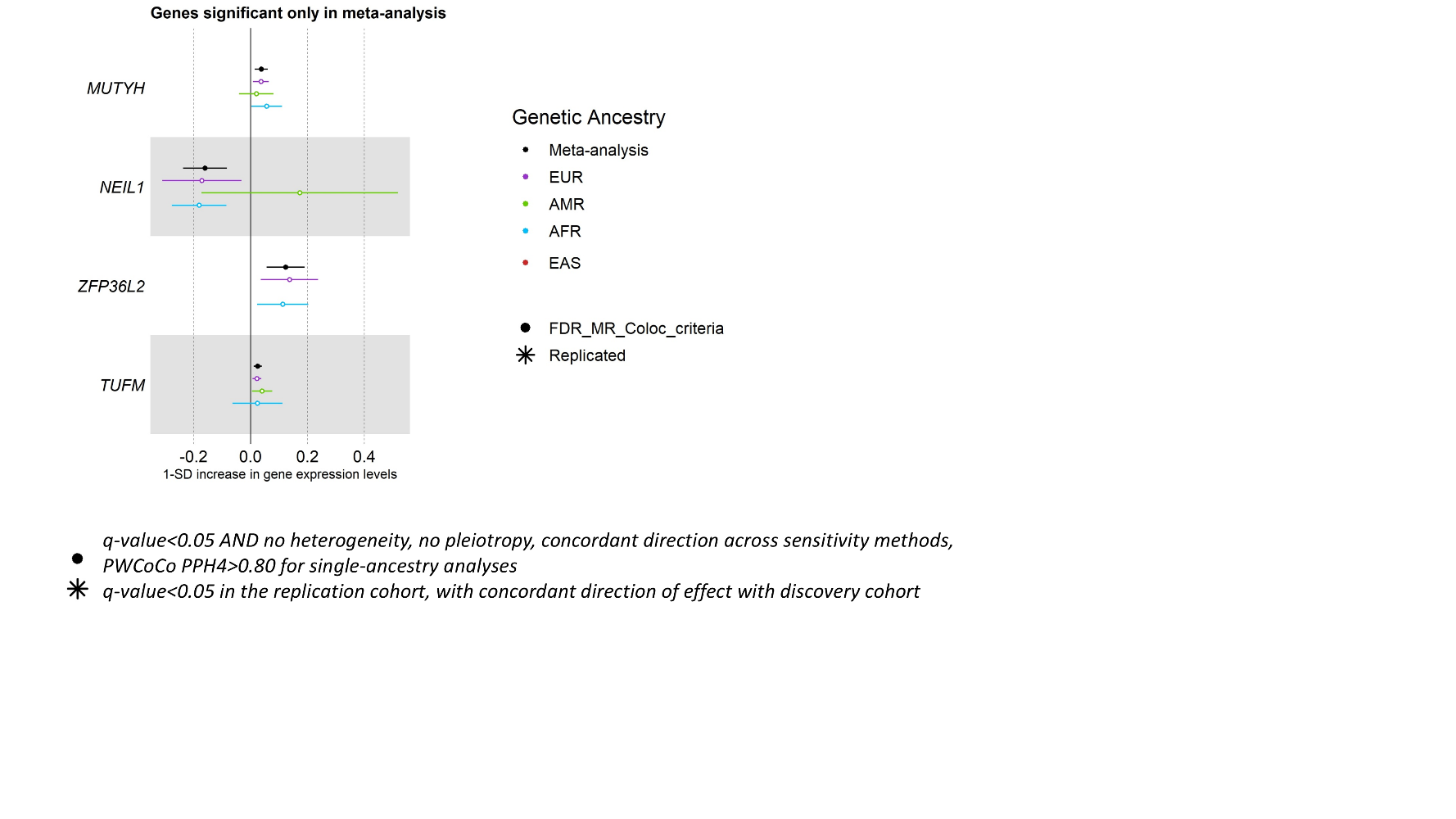

***Supplemental Figure 1****: Forest plots of the causal effects identified in the blood eQTL MR analyses for genes detected only in the meta-analysis but not in single-ancestry MR analyses. Causal estimates from the single-ancestry MR in the discovery cohorts (matched genetic ancestry group between the exposure and the outcome) are also represented. Filled dots represent causal estimates from MR analyses that have a q-value<0.05, and (1) pass the sensitivity criteria and show evidence of colocalization (PPH4>0.8) in single-ancestry analyses, or (2) present nominal significance and meet criteria (1) in at least one cohort entering the meta-analysis. Genes and proteins with causal effects identified in single-ancestry analyses and replicated in independent cohorts from the same genetic ancestry group are denoted with a star. We report causal estimates as odds ratios (OR) for T2D per standard deviation (SD) change in genetically predicted gene expression or protein levels.*

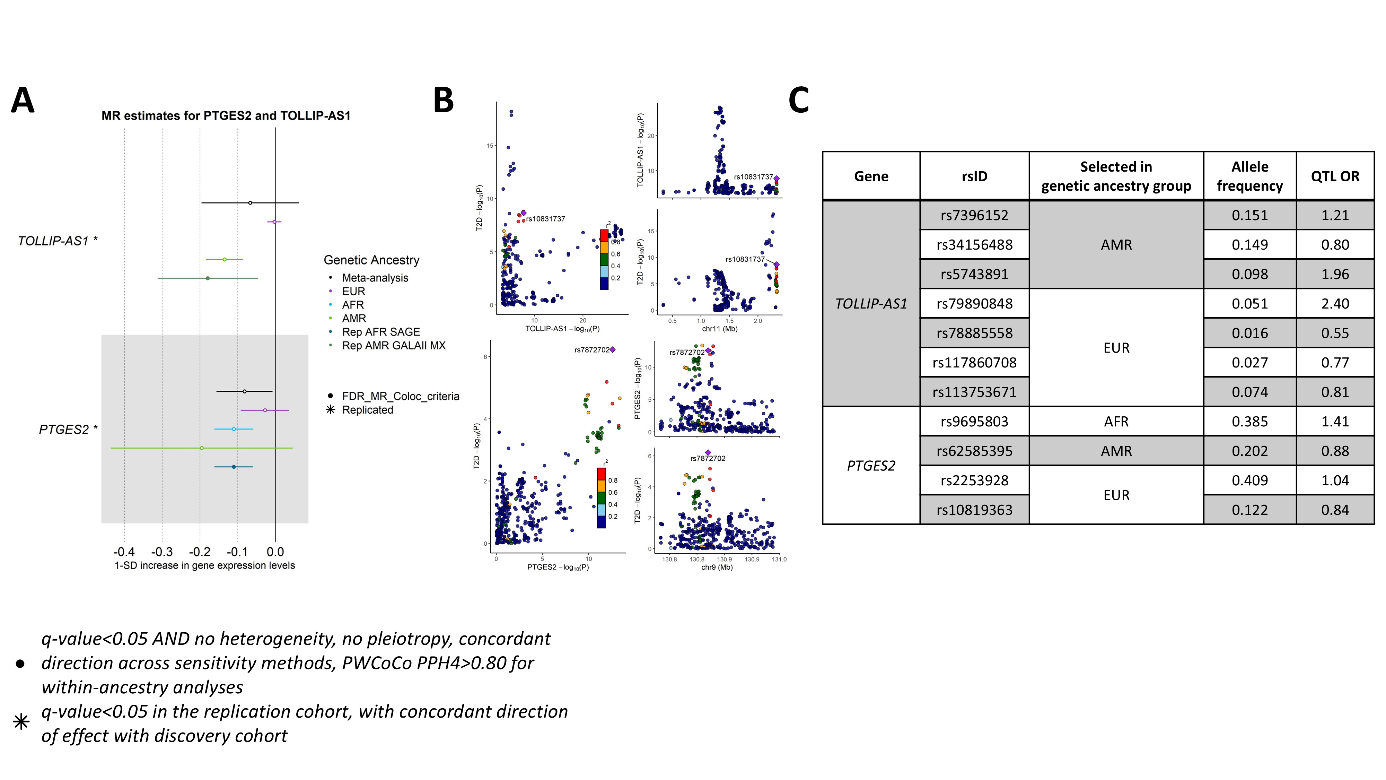

***Supplemental Figure 2:*** *Causal estimates and frequency and effect sizes of IVs on TOLLIP-AS1 and PTGES2 (A) Forest plots of causal estimates for TOLLIP-AS1 and PTGES2 from the eQTL MR analyses. Causal estimates from the single-ancestry MR in the discovery cohorts (matched genetic ancestry group between the exposure and the outcome) are also represented. Filled dots represent causal estimates from MR analyses that have a q-value<0.05, and (1) pass the sensitivity criteria and show evidence of colocalization (PPH4>0.8) in single-ancestry analyses, or (2) present nominal significance and meet criteria (1) in at least one cohort entering the meta-analysis. Genes and proteins with causal effects identified in single-ancestry analyses and replicated in independent cohorts from the same genetic ancestry group are denoted with a star. We report causal estimates as odds ratios (OR) for T2D per standard deviation (SD) change in genetically predicted gene expression or protein levels. (B) LocusCompare and LocusZoom for the two genes demonstrating colocalization evidence. (C) Allele frequency of the IVs selected for the two genes, and their association with the corresponding gene expression levels.*

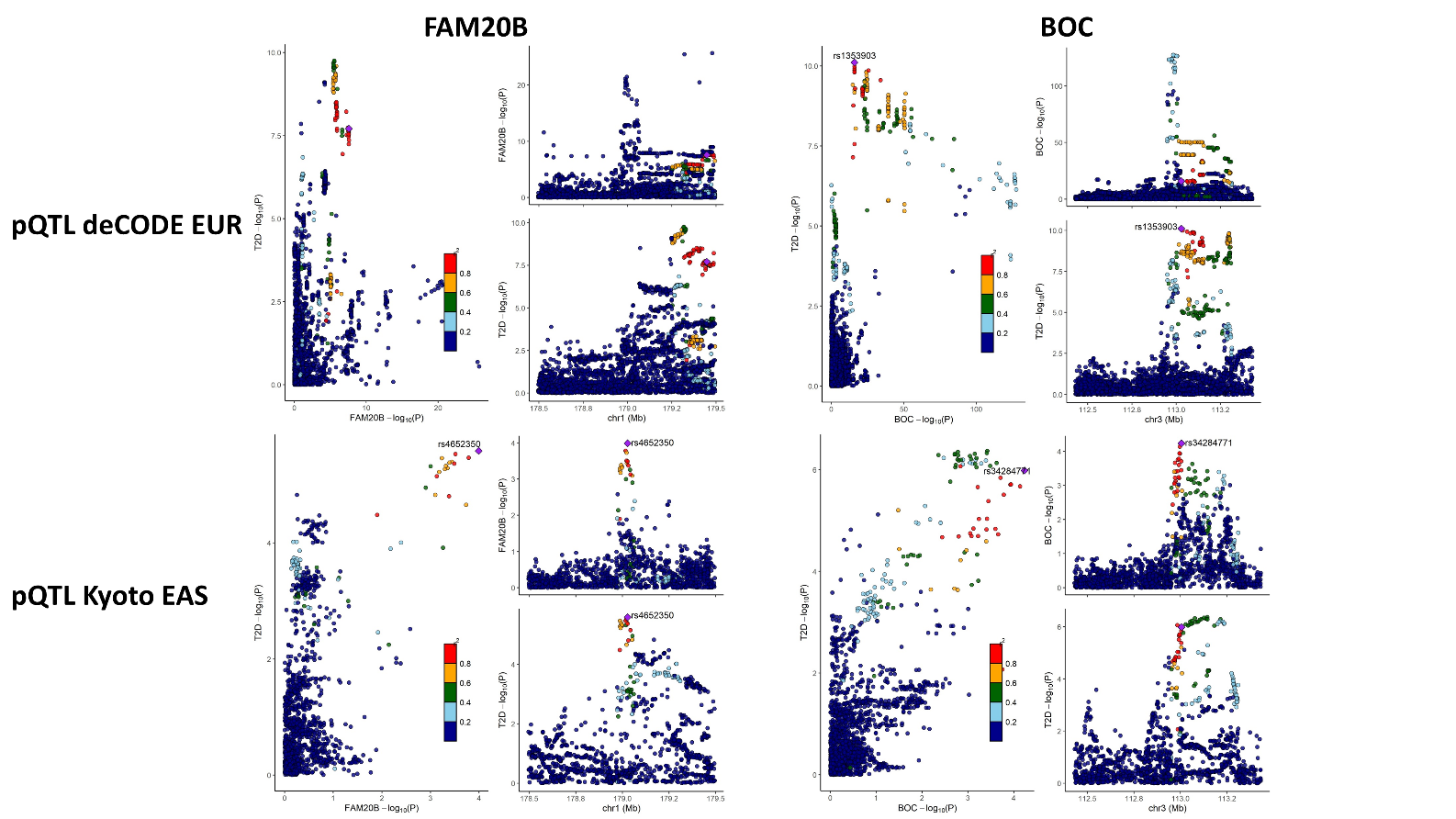

***Supplementary Figure 3:*** *LocusCompare and LocusZoom for FAM20B and BOC, two proteins with significant causal effects detected only in EAS. Results are displayed in EUR (top panel) and in EAS (bottom panel).*

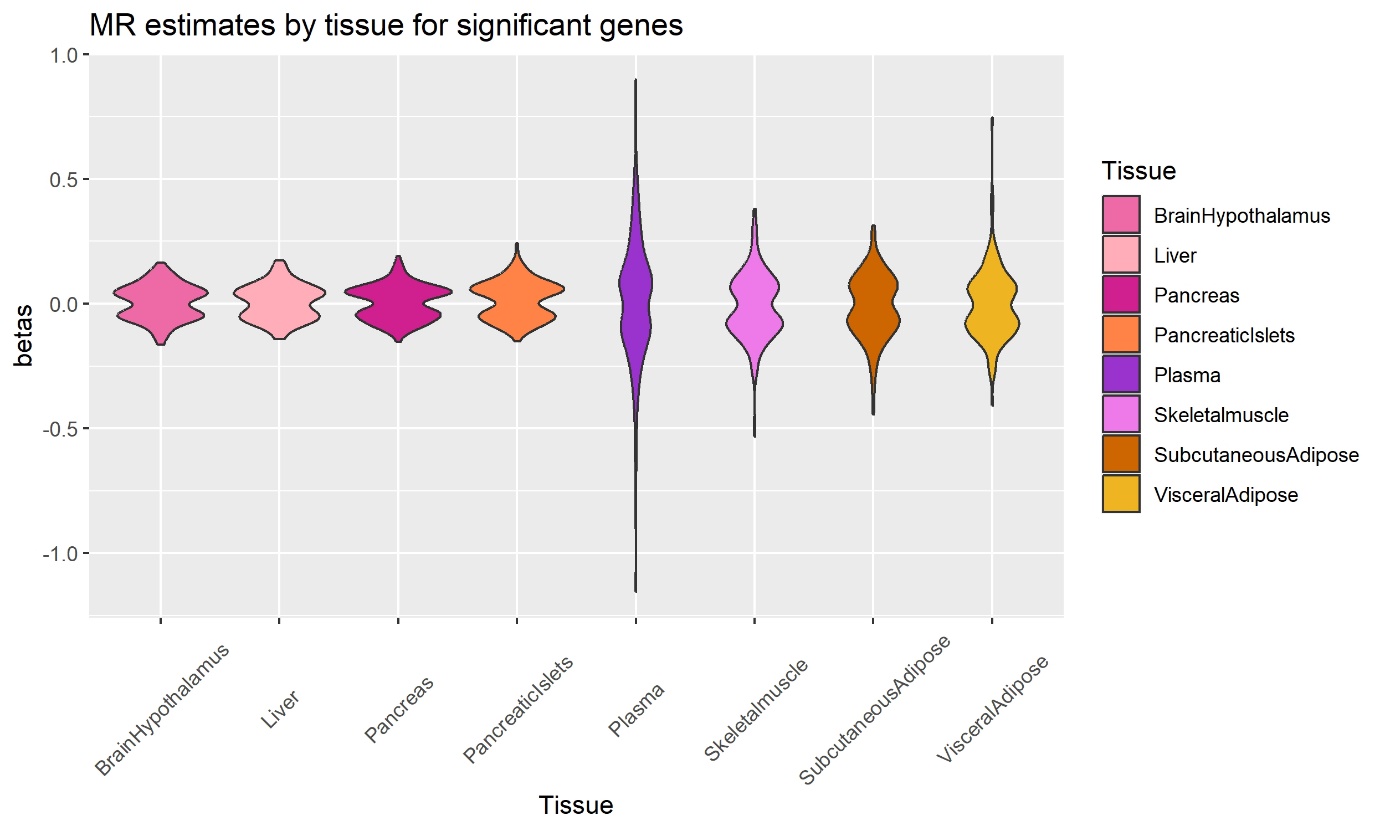

***Supplementary Figure 4:*** *Distribution of significant causal effect estimates according to the tissue of the eQTL dataset.*

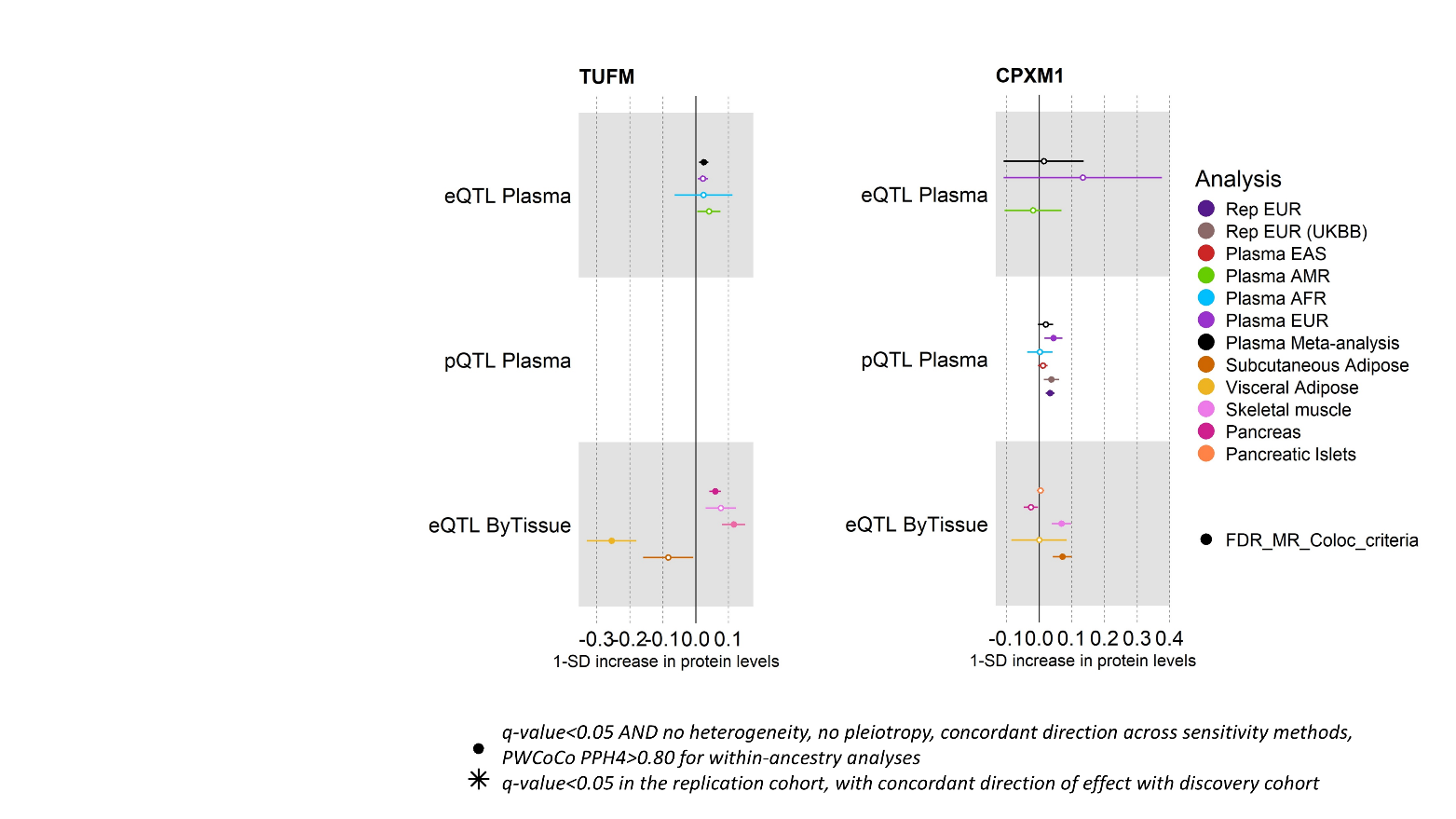

***Supplementary Figure 5:*** *Forest plots of causal estimates for TUFM and CPXM1 from all the MR analyses. Causal estimates from the single-ancestry MR in the discovery cohorts (matched genetic ancestry group between the exposure and the outcome) are also represented. Filled dots represent causal estimates from MR analyses that have a q-value<0.05, and (1) pass the sensitivity criteria and show evidence of colocalization (PPH4>0.8) in single-ancestry analyses, or (2) present nominal significance and meet criteria (1) in at least one cohort entering the meta-analysis. Genes and proteins with causal effects identified in single-ancestry analyses and replicated in independent cohorts from the same genetic ancestry group are denoted with a star. We report causal estimates as odds ratios (OR) for T2D per standard deviation (SD) change in genetically predicted gene expression or protein levels.*

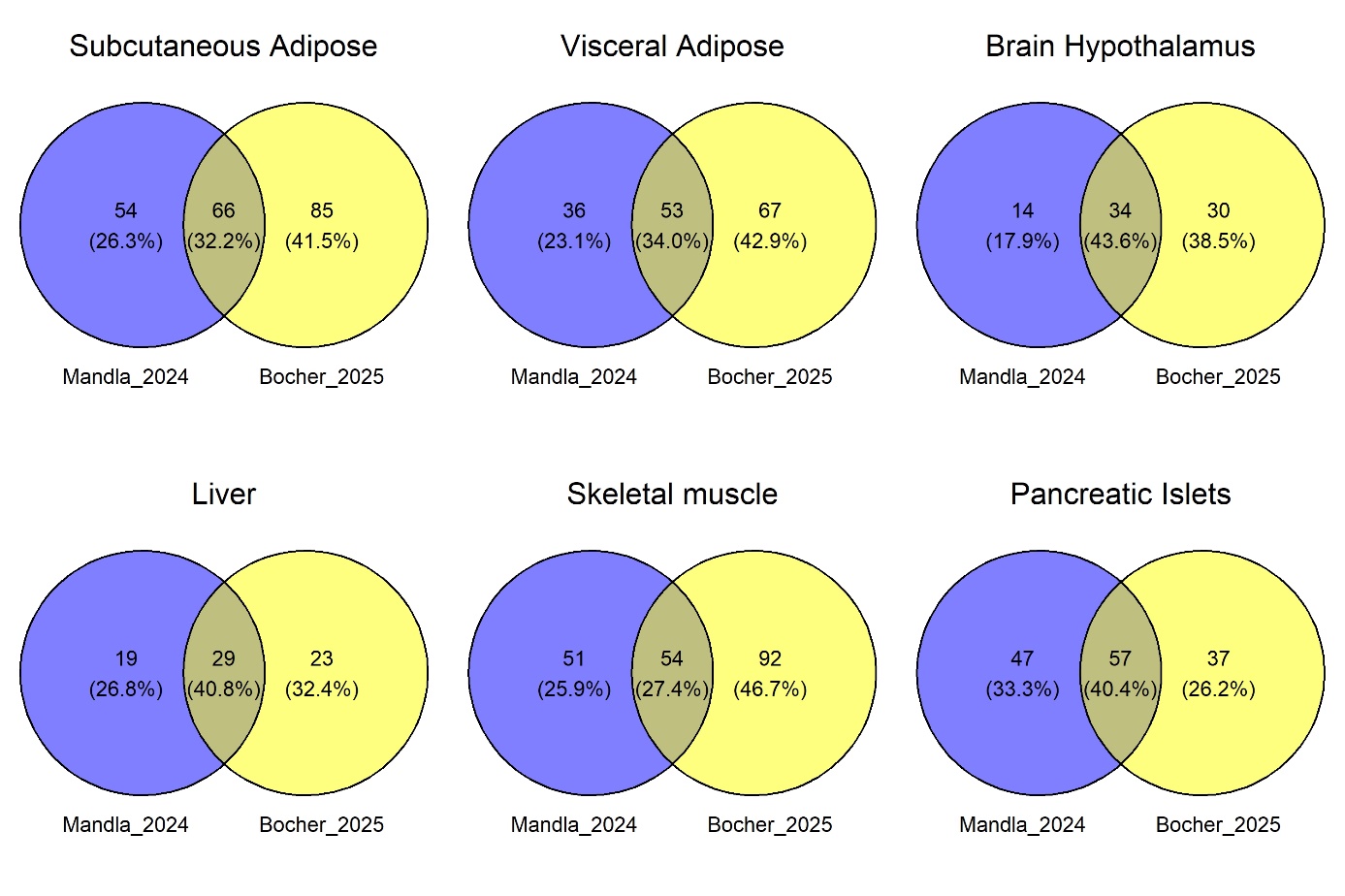

***Supplementary Figure 6:*** *Venn Diagrams representing the overlap of genes identified in the previous colocalization study from Mandla et al. 2024, and in the present study (MR+colocalization using PWCoCo). Comparisons are indicated for six tissues which used the same QTL datasets in both studies.*

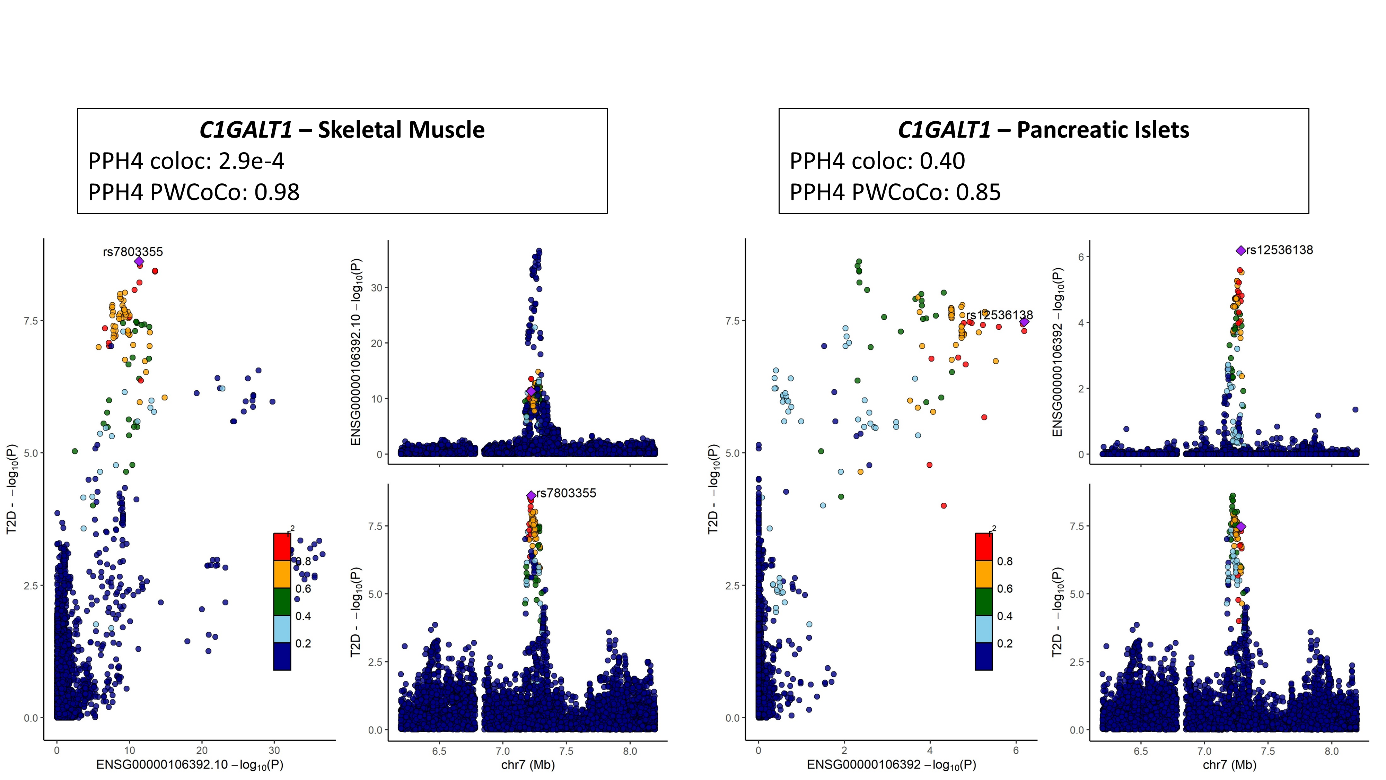

***Supplementary Figure 7:*** *LocusCompare and LocusZoom for C1GALT1 in Skeletal Muscle and in Pancreatic Islets. PPH4 obtained with the coloc approach (from Mandla et al. 2024) and with the PWCoCo approach (the present study) are indicated.*
